# Estimating the gap between clinical cholera and true community infections: findings from an integrated surveillance study in an endemic region of Bangladesh

**DOI:** 10.1101/2023.07.18.23292836

**Authors:** Sonia Hegde, Ashraf Islam Khan, Javier Perez-Saez, Ishtiakul Islam Khan, Juan Dent Hulse, Md Taufiqul Islam, Zahid Hasan Khan, Shakeel Ahmed, Taner Bertuna, Mamunur Rashid, Rumuna Rashid, Md Zakir Hossain, Tahmina Shirin, Kirsten Wiens, Emily S. Gurley, Taufiqur Rahman Bhuiyan, Firdausi Qadri, Andrew S. Azman

## Abstract

Our understanding of cholera transmission and burden largely rely on clinic-based surveillance, which can obscure trends, bias burden estimates and limit the impact of targeted cholera-prevention measures. Serologic surveillance provides a complementary approach to monitoring infections, though the link between serologically-derived infections and medically-attended disease – shaped by immunological, behavioral, and clinical factors – remains poorly understood. We unravel this cascade in a cholera-endemic Bangladeshi community by integrating clinic-based surveillance, healthcare seeking, and longitudinal serological data through statistical modeling. We found >50% of the study population had a *V. cholerae* O1 infection annually, and infection timing was not consistently correlated with reported cases. Four in 2,340 infections resulted in symptoms, only one of which was reported through the surveillance system. These results provide new insights into cholera transmission dynamics and burden in the epicenter of the 7^th^ cholera pandemic and provide a framework to synthesize serological and clinical surveillance data.

## Introduction

More than half a century into the 7th cholera pandemic, *Vibrio cholerae* El Tor O1 continue to ravage communities lacking access to safe water and insanitation. In contrast to the few official cholera cases reported to the World Health Organization each year from South Asia, this region likely bears a significant portion of the global burden of cholera and serves as the source of genetic diversity within the 7th pandemic lineage ^1–3^.

While past surveillance efforts have provided glimpses into the burden of cholera in South Asia, their scope has been limited due to non-exhaustive testing of acute watery diarrhea cases (suspected cholera), which could result from a number of other related pathogens, and a focus on only medically-attended cases ^4–7^.

Viewing cholera through the lens of passive surveillance systems alone may lead to severe underestimates of the incidence and a skewed understanding of the transmission dynamics of and population immunity related to pandemic *V. cholerae* O1. This understanding is critical for designing tailored interventions to curb transmission and reduce the burden of disease, especially in the posited cholera epicenter of South Asia.

Historical estimates of cholera seroincidence from South Asia have revealed a wide gap between infections – defined by an immunologic boost of *V. cholerae* O1 specific antibodies – and the number of cholera cases detected through clinic-based surveillance ranging from close to 1 case per infection to well over 100 ^8–10^. More recent analyses from a national serosurvey in Bangladesh suggested that 1 in 5 people have at least one infection each year with *V. cholerae* O1, yet fewer than 1,000 cases have been reported to the WHO annually over the past decade ^11,12^. Though both serologically inferred infections and clinical cases provide important clues about the true burden of disease, each data type has its own limitations and biases, and methods to integrate both types of data and translate one derived metric to another (i.e., infections to medically attended cases, and vice versa) are lacking.

Only a fraction of infections appear as confirmed cases within a facility-based surveillance system which can be due to a suite of host and pathogen factors (Figure 1). Serologically-inferred infections represent exposures to *V. cholerae* O1 leading to a boost in antibodies, but only a fraction of these lead to clinical disease, which can partially be explained by differences in exposure routes, inoculum size, previous exposures to *V. cholerae* O1 and circulating strains ^8^. Further, upon experiencing a symptomatic *V. cholerae* O1 infection, only a portion of individuals will seek care at formal medical facilities based on disease severity and access to care. While diarrhea-related care seeking data for children less than five years of age are abundant, largely through the Demographic and Health Surveys (DHS) ^13,14^, such data for older children and adults are scant, thereby limiting methods for translating estimates of infection into estimates of burden of disease. Passive clinic-based surveillance is most commonly employed for tracking cholera mainly through the symptom-based classification of suspected cholera, however, laboratory confirmation is rare and the suspected cholera case definition is known to be highly non-specific ^15^. A recent review suggested that, on average, less than half of clinically suspected cholera cases captured by surveillance are true infections from *V cholerae* O1, but this proportion varied widely over time and space ^16^. Understanding the pathway from infections with *V. cholerae* O1 to confirmed and reported clinical cholera is crucial for interpreting surveillance data, making inferences about transmission dynamics and disease burden, and ultimately improving our ability to target resources in the fight against cholera.

**Figure 1.**
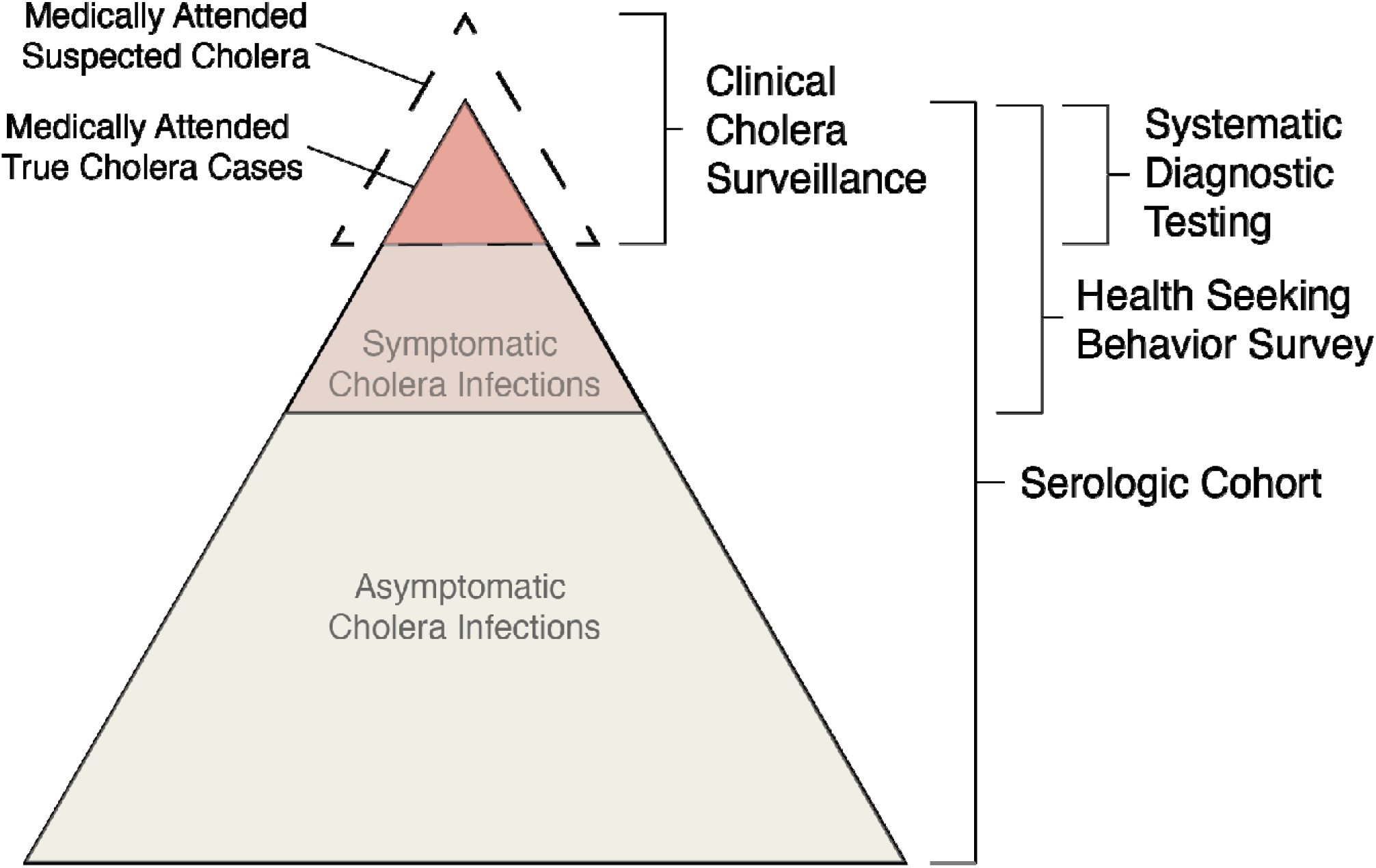
Overview of conceptual model of the continuum between infections and medically attended cholera. Key data sources used in the model are indicated on the right.

Here, we combine data from a longitudinal serologic cohort, enhanced clinical and laboratory surveillance, and a health seeking behavior survey to elucidate the pathway between exposure to these bacteria and clinical disease, and estimate the true incidence of infections and symptomatic cholera disease in an endemic sub-district of Bangladesh ^7,11^. Refining our understanding of the links between *V. cholerae* O1 infections in the community and medically attended clinical cholera is key to enhancing our ability to interpret data from passive clinical surveillance and serosurveys in South Asia and elsewhere.

## Results

### Medically attended suspected cholera cases are reported throughout the year

Through enhanced clinical surveillance from January 24, 2021 to February 13, 2022, we detected 2,176 suspected cholera cases (defined as those ≥1 years old experiencing 3 or more watery, non-bloody stools in the last 24 hours) at the two primary sites treating diarrhea cases in the Sitakunda subdistrict, the geographic focus of our analyses (Figure S1). Thirty-four percent of suspected cases were less than 5 years of age and forty-six percent (998/2176) of suspected cases were from or spent the last seven days in Sitakunda. Nearly all (99.6%) of the suspected cases from Sitakunda were admitted to the health facility and experienced some (96%) or severe (3%) dehydration (Table 1); there were no reported deaths. The annual incidence rate of suspected cholera cases in Sitakunda alone was 2.1 per 1,000, including 10.5 per 1,000 among those 1-4 years old, 1.2 per 1,000 among those 5-64 years old and 1.9 per 1,000 among those 65 years and older.

**Table 1.**
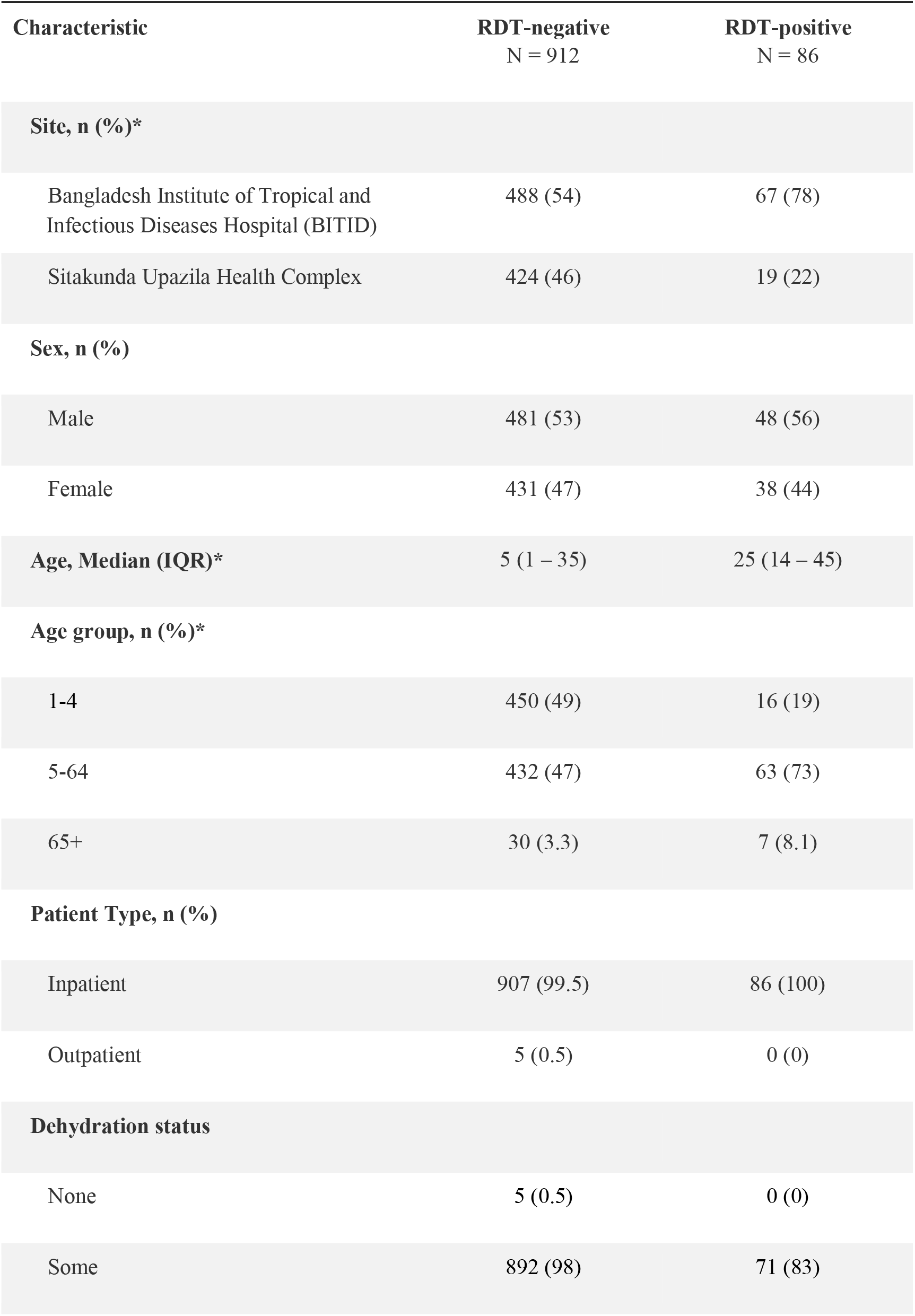
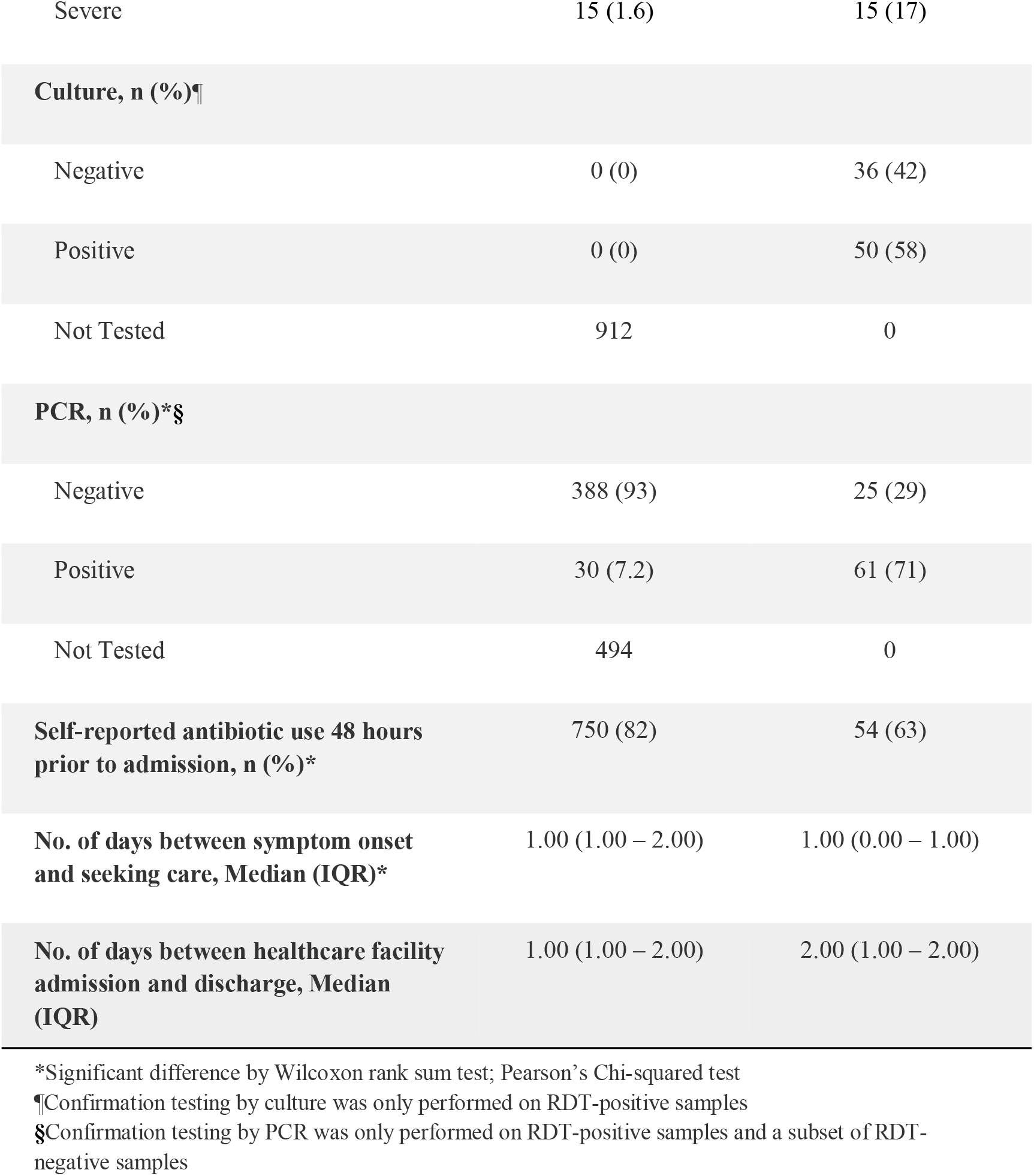
Overview of suspected cases by RDT status for participants enrolled into clinical surveillance from January 24, 2021 to February 13, 2022, the end of the last serosurvey, that resided in the Sitakunda sub-district.

### True medically attended cholera cases reveal different temporal and age-specific trends from suspected cases

As the suspected case definition is known to have low specificity for infections with *V. cholerae* O1^15^, we performed rapid diagnostic tests on all suspected cholera cases and PCR and/or culture on a subset. After taking into account the performance of the array of diagnostic tests used (Table 1), we reconstructed the true weekly incidence of medically attended cholera cases, which displayed a distinct temporal signature with the majority of cases (54%) confined to just a four-month period (April to July 2021, Figure 2). The incidence rate of medically attended cholera (0.2 per 1,000 per year; 95% Credible Interval [CrI] 0.2-0.3) was roughly one-tenth of the observed suspected cholera incidence rate, with children 1-4 and those over 65 years old having 2-5 times the medically-attended incidence rates as others (Table 3). This implies that for every 10 (95% CrI 7- 10) suspected cholera cases at facilities in Sitakunda, one of these is caused by *V. cholerae* O1, with strong temporal and age-specific heterogeneity, likely due to the incidence of other seasonal pathogens (Table S2)^17,18^.

**Figure 2.**
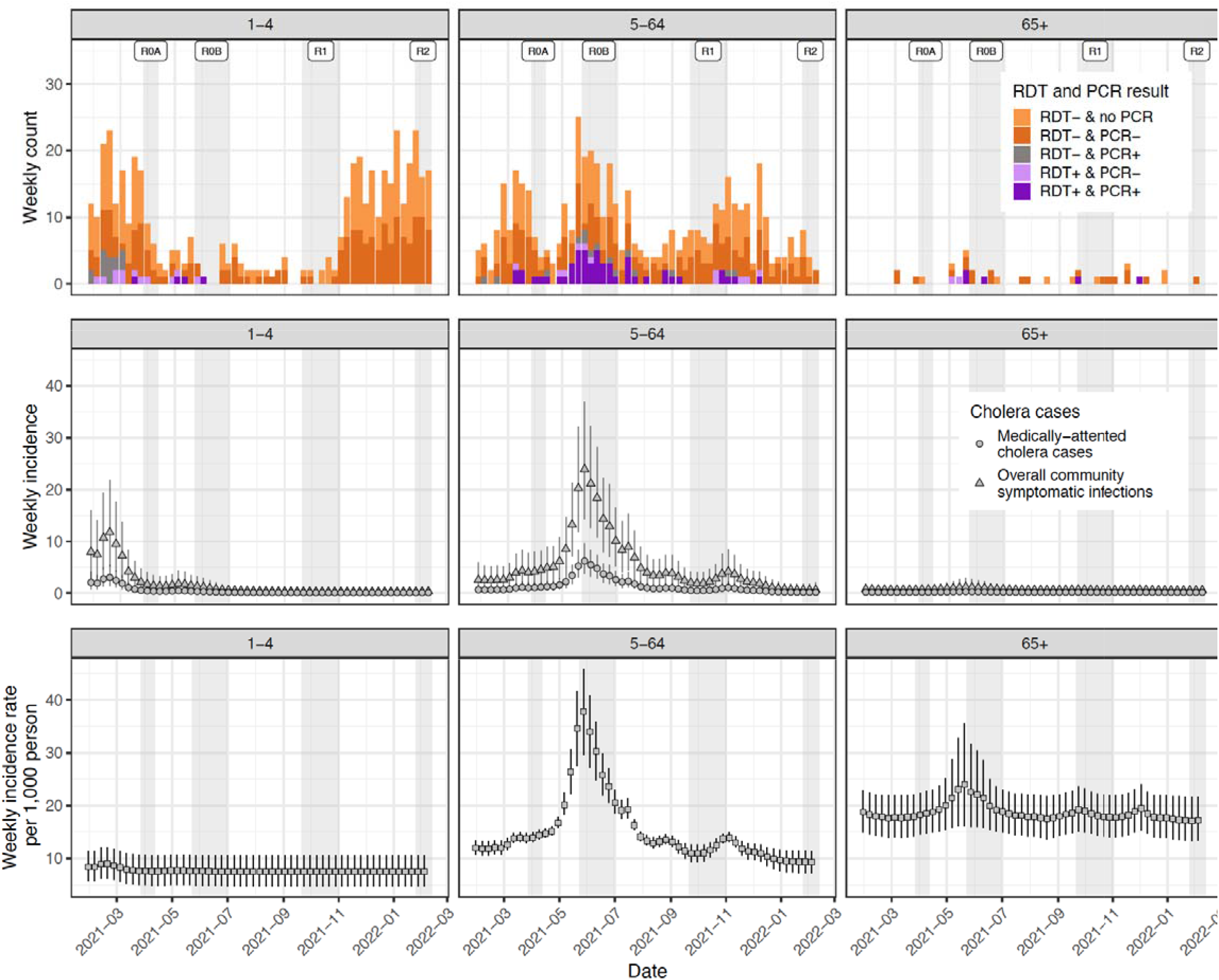
Weekly suspected cases (top row), symptomatic infections (middle row), and exposures/infections (bottom row) by age group. The top row illustrates the weekly number of suspected cases colored by RDT and PCR results (culture results not shown). All suspected cases were tested by RDT but only positives and around half of negatives were tested by PCR. The middle row illustrates weekly estimates of the number of true cholera cases seeking care at study facilities (circles) with triangles representing the estimates of the true number of symptomatic cases both in facilities and the community. The bottom row represents estimates of the number of weekly infections that elicit an immunologic boost, as inferred from longitudinal vibriocidal titers. Vertical bars across the middle and bottom rows represent the 95% credible intervals.

### Health seeking behavior masks the true burden of symptomatic cholera infections in the community

Not all people will seek health care for diarrhea and among those that do, only a fraction will report to facilities that are part of official disease surveillance systems ^19^. To understand care seeking behaviors for cholera-like symptoms, we conducted a survey of 2,481 individuals from 580 households (see Methods) representative of the catchment area of clinical surveillance and asked each person about their health care seeking preferences. The survey included 53% females and had a similar demographic profile to the national population with notable under sampling of young children (Table 2, Figure S2). When asked whether they would seek care for acute watery diarrhea with some dehydration, 86% (N=2,128) indicated they would, with the likelihood decreasing by age (Table S1). Of the participants that said they would seek care, over half (57.6%) said they would seek care at a pharmacy, and 30% said they would seek care at one of the two official clinical surveillance facilities in Sitakunda. Overall, we estimate that for every 4 (95% CrI 3.7-4.1) individuals who are symptomatic, including with diarrhea, only one would seek care at the official cholera surveillance facilities in Sitakunda and be captured as a suspected cholera case, with no significant differences by age (Figure 3).

**Figure 3.**
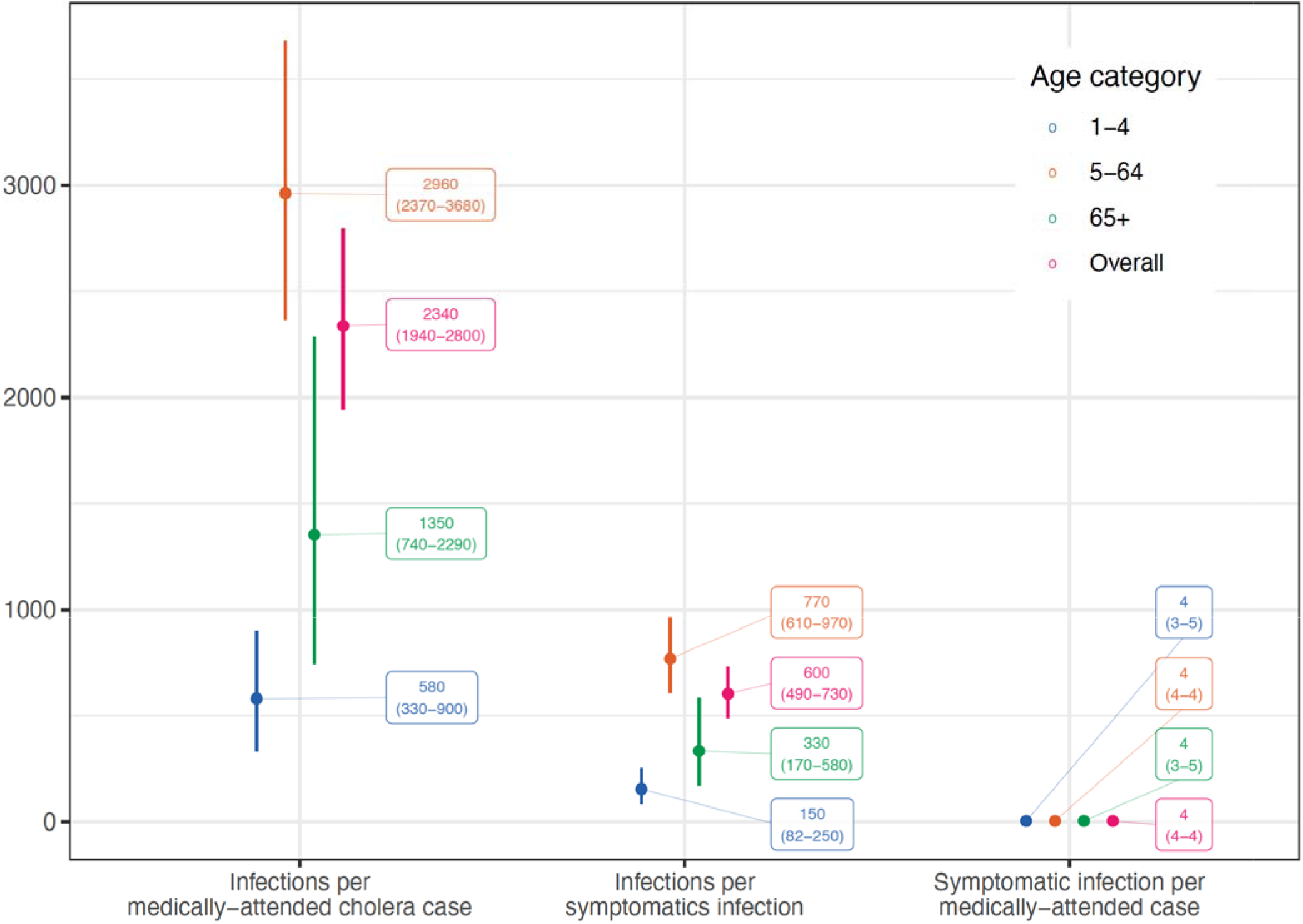
Estimates of ratios along the continuum between infections and reported medically-attended cholera cases by age group (color).

**Table 2.**
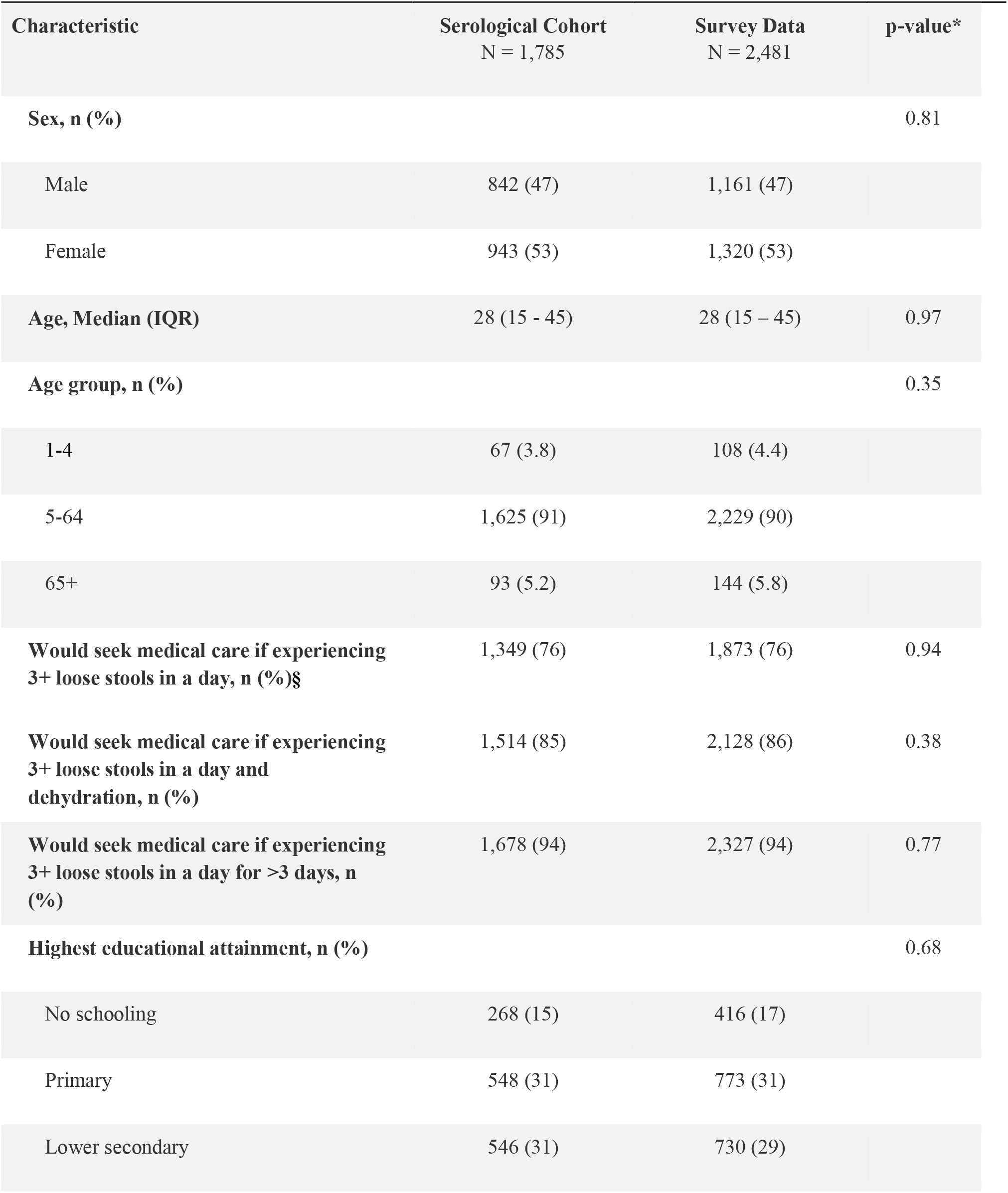
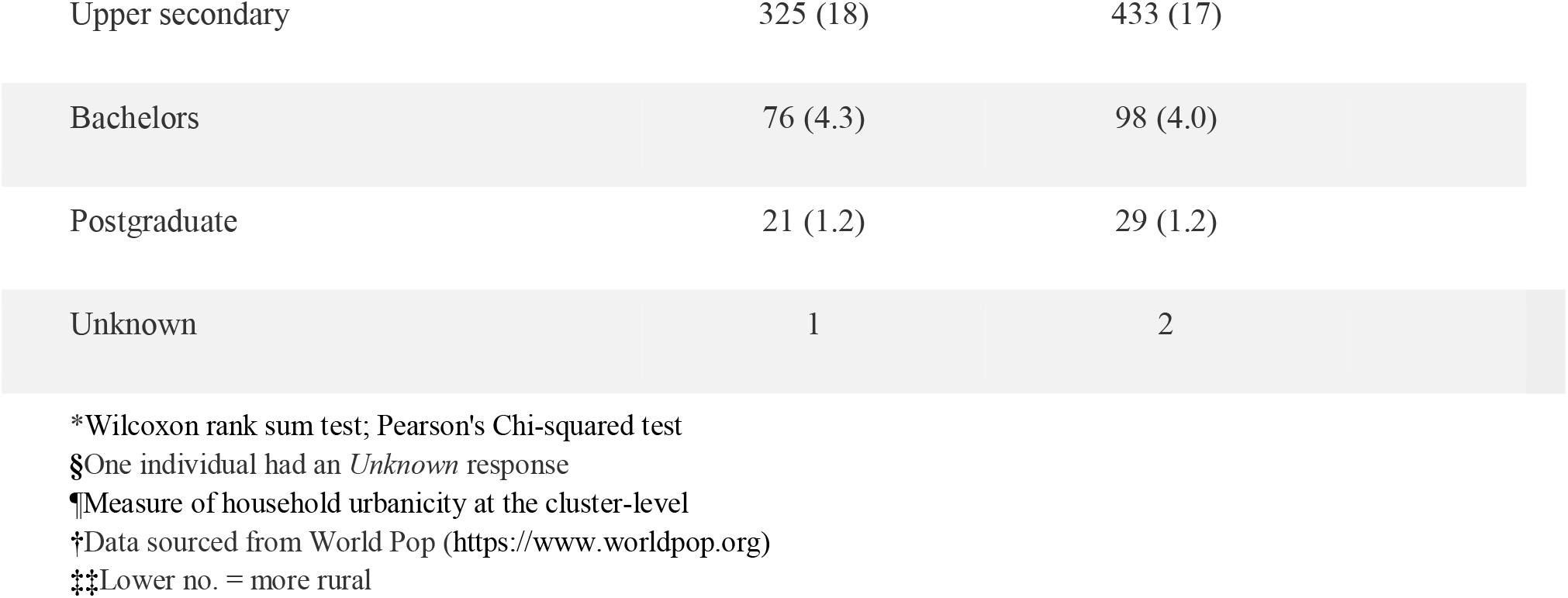
Overview of the participants enrolled into the serosurvey. These data are separated by those who were enrolled and provided serum samples for all three rounds of the serosurvey (N=1,785; Serological Data) and those who were enrolled and provided survey data across the entire study period (N=2,481; Survey Data). There are no statistically significant differences among the two survey populations used by the following demographic characteristics.

| Characteristic | Serological Cohort<br>N = 1,785 | Survey Data<br>N = 2,481 | p-value* |
| --- | --- | --- | --- |
| **Sex, n (%)** |  |  | 0.81 |
| Male | 842 (47) | 1,161 (47) |  |
| Female | 943 (53) | 1,320 (53) |  |
| **Age, Median (IQR)** | 28 (15 - 45) | 28 (15 – 45) | 0.97 |
| **Age group, n (%)** |  |  | 0.35 |
| 1-4 | 67 (3.8) | 108 (4.4) |  |
| 5-64 | 1,625 (91) | 2,229 (90) |  |
| 65+ | 93 (5.2) | 144 (5.8) |  |
| **Would seek medical care if experiencing 3+ loose stools in a day, n (%)§** | 1,349 (76) | 1,873 (76) | 0.94 |
| **Would seek medical care if experiencing 3+ loose stools in a day and dehydration, n (%)** | 1,514 (85) | 2,128 (86) | 0.38 |
| **Would seek medical care if experiencing 3+ loose stools in a day for >3 days, n (%)** | 1,678 (94) | 2,327 (94) | 0.77 |
| **Highest educational attainment, n (%)** |  |  | 0.68 |
| No schooling | 268 (15) | 416 (17) |  |
| Primary | 548 (31) | 773 (31) |  |
| Lower secondary | 546 (31) | 730 (29) |  |

|  |  |  |
| --- | --- | --- |
| Upper secondary | 325 (18) | 433 (17) |
| Bachelors | 76 (4.3) | 98 (4.0) |
| Postgraduate | 21 (1.2) | 29 (1.2) |
| Unknown | 1 | 2 |

\*Wilcoxon rank sum test; Pearson's Chi-squared test
§One individual had an *Unknown* response
¶Measure of household urbanicity at the cluster-level
‡Data sourced from World Pop (https://www.worldpop.org)
‡‡Lower no. = more rural

### Infections with V. cholerae O1 are frequent though few lead to symptomatic disease

Though *V. cholerae* O1 infections manifest across the spectrum of clinical disease severity, the proportion of infections leading to symptomatic disease is unclear. To understand the incidence of infections, independent of clinical manifestations, we followed participants of the representative health-seeking survey over the course of one year and collected three blood draws per person (n=1,785 participated in all three visits; Figure 1, Table 2). We measured vibriocidal titers, an established marker of recent infection, for each sample following standard methods ^20,21^. Given that vibriocidal titers decay within the time between study visits (half-life of 74-201 days ^22^; time between study visits was 87-214 days), we developed a Bayesian approach to reconstruct the unobserved infection histories of each participant in our serologic cohort taking into account both antibody kinetics and measurement error (see Methods and Supplement S4).

Combining the serologic trajectories with the reconstructed incidence of symptomatic cholera, we estimated an annual *V. cholerae* O1 infection incidence rate of 525 per 1,000 population (95% CrI 508-550), with incidence increasing by age group (Table 3). Those 1-4 years old had an infection incidence of 296 per 1,000 (95% CrI 199-395), which was notably lower than other age groups. For every 600 infections (95% CrI 490-730), we estimated that only one infection will result in symptomatic disease. Young children and the elderly had a two to five times higher probability of symptomatic disease given infection than the rest of the population, suggestive of differences in exposure routes, prior immunity, and/or a lower infectious dose threshold needed to cause clinically relevant disease. The incidence rate of symptomatic cholera, including both those that are medically attended and not, was 0.9 per 1,000 per year (95% CrI 0.7-1.0) with young children and the elderly having twice the incidence rate compared to others (Table 3).

**Table 3.** Estimates of annualized incidence rates per 1,000 population.

|  | ≥1-4 years | 5-64 years | >65 years | Overall |
| --- | --- | --- | --- | --- |
| **Healthcare facility incidence rates (per 1,000 population)** |  |  |  |  |
| Suspected cholera incidence | 10.5 | 1.2 | 1.9 | 2.1 |
| RDT-positive incidence | 0.4 | 0.2 | 0.4 | 0.2 |
| Medically-attended cholera incidence | 0.5<br>(0.4-0.7) | 0.2<br>(0.1-0.2) | 0.5<br>(0.3-0.8) | 0.2<br>(0.2-0.3) |
| **Community incidence rates (per 1,000 population)** |  |  |  |  |
| Symptomatic cholera incidence<br>(community + clinics) | 2.1<br>(1.3-3.0) | 0.7<br>(0.6-0.9) | 2.0<br>(1.0-3.4) | 0.9<br>(0.7-1.0) |
| Incidence of infections<br>(community) | 295.6<br>(199.1-395.3) | 530.5<br>(513.0-556.4) | 599.8<br>(513.6-686.4) | 525.3<br>(508.3-550.3) |

### Clinic-based cholera surveillance leads to a biased view of V. cholerae O1 infections in the community

Viewing cholera transmission through only the lens of clinic-based surveillance can lead to biases in both the scale and temporal nature of transmission. Our estimates imply that for every 2,340 (95% CrI 1,940- 2,800) infections in the community, only one ended up as a medically attended cholera case captured by the surveillance system, and if cholera confirmation was done with traditional microbiological culture, this gap would grow even wider due to imperfect test performance (3,020 infections per medically-attended cholera case; 95% CrI 2,480-3,660).

The relationship between infections in the community and observed clinical cholera cases was not constant through time. We initially conceptualized that the rates of clinical cholera and infections were proportional throughout the study period in our Bayesian model of infection incidence. However, we found strong statistical support (LOO-PSIS difference of 78.7, SE = 19.0) for an alternative model where seroincidence rates were composed of two components: one proportional to clinical cholera incidence rates and the other a constant hazard throughout the period independent of observed clinical cases, which was ultimately used in our primary analyses. This model formation, which served as the basis for our primary results, suggests that even at times with no medically attended cholera cases, more than four thousand infections (weekly range: 4,352-16,154, annualized weekly incidence rate 9.8-36.6 per 1,000 people per year) occur each week in Sitakunda.

## Discussion

Through joint inference on data from enhanced clinical and laboratory surveillance, a longitudinal serologic cohort, and a healthcare seeking behavior survey, we illustrate a landscape of regular immunologic exposures to *V. cholerae* O1 where resulting cholera disease is the exception rather than the rule. We estimated that only 4 in 2,340 infections will result in symptomatic infections and among these, only one will seek care and ultimately be captured by the surveillance system. Passive clinic-based surveillance, the lens through which cholera is typically viewed in most of the world, captures only a fraction of symptomatic cases and obscures our understanding of the dynamics and magnitude of *V. cholerae* O1 infections. Our results shed new light on the continuum between infections and medically attended clinical disease in endemic populations and provide robust estimates of the true burden of cholera.

While several investigators and decision-making bodies have attempted to estimate the true burden of symptomatic cholera, they have primarily used facility-based suspected case data, often without laboratory confirmation. Most studies have neither adjusted for health seeking behaviors, diagnostic test performance (when used) nor severity of infection, possibly resulting in biased estimates of risk and poor allocation of resources. Perhaps the most widely used estimate of cholera incidence in Bangladesh, for several policy and planning documents, was based on an extrapolation of medically-attended cholera incidence measured in 2005 in the neighboring area of Kolkata, India (∼1.64 per 1,000 people) ^23^. Another study conducted in six sentinel sites around Bangladesh from 2010-2011, which attempted to estimate the incidence of severe cholera by use of hospital admissions data and a care-seeking survey, found that incidence rates ranged across sites from 0.3 to 4.9 per 1,000 per year and care seeking probabilities consistent with those estimated in our study ^24^. Unlike previous studies, we incorporated serologically-derived incidence data and accounted for diagnostic test performance in our estimation, and approximated the true burden of cholera in this endemic setting to be 0.9 symptomatic infections per 1,000 per year. Though our estimate falls within the lower range of historical estimates, we cannot reliably attribute differences to true changes in incidence given the prior use of alternate, less sensitive methods.

Our approach allowed us to estimate and partition the overall incidence of *V. cholerae* O1 infections, those that result in symptoms and eventually care-seeking, and those that remain subclinical (Figure 1). Far greater than previous estimates from cross-sectional serological data alone (20%, 95% CrI 104-352; for all of Chattogram District ^11^), we estimated that at least 50% of the Sitakunda population is infected with *V. cholerae* O1 annually, and that >99% of these infections are subclinical. Such a high-level of infection incidence has only been documented in outbreaks in previously naive populations, similar to after the initial introduction of cholera in Haiti in 2010 ^25^, and thus has been attributed to a lack of pre-existing immunity. The high proportion of subclinical infections observed in Sitakunda, however, could represent transient infections or exposures that do not result in gut colonization or bacterial shedding and/or asymptomatic or less severe infections that result in secondary transmission. Both of these plausible explanations have implications for our understanding of endemic cholera transmission dynamics. The observed high incidence rate in Sitakunda combined with our understanding of the reproductive number of cholera (range 1-3, ^26–29^) and the biannual seasonal cholera peaks ^7^ are consistent with the hypothesis that not all infections lead to the same magnitude and duration of protection against clinical disease. Previous modeling work implied that mild and asymptomatic infections ^30^ or inadequate levels of cross-protection between serotypes ^31^ may lead to faster immune waning, resulting in endemic areas bearing a consistently high infection incidence, but strong empirical evidence is still lacking. Despite the apparent disconnect between infection and subsequent protection, which may be explained by heterogeneity in inoculum size, immune responses and/or infection histories, our study highlights potential hypotheses and corroborates these as targets for future investigations.

By inferring time-varying infections, our results contribute to our understanding of the different pathways through which cholera transmission occurs. Two views on the dominant routes of *V. cholerae* infections leading to clinical disease have emerged over the past 30 years: the first suggesting that infections occur through exposures to *V. cholerae* in the aquatic environment and the second that infections are through more proximal exposures to infected individuals, often via short-term conduits like food or water ^32,33^. We found robust statistical support that infections in Sitakunda could be explained by two components, one linked to the number of cases observed at health facilities and the other a constant hazard from an unknown source. This finding may be consistent with the dual roles of frequent low-dose exposures to environmental *V. cholerae* (O1 and other serogroups) and seasonal exposures to higher doses of bacteria, potentially of the hyperinfectious phenotype ^34^, from symptomatic individuals. As the Bay of Bengal forms the entire western border of Sitakunda, frequent exposures to aquatic flora and fauna, which have been associated with *V. cholerae* infection, is plausible ^35^. A well-documented dose-response relationship exists as higher doses of *V. cholerae* O1 bacteria lead to more severe disease ^36^, and this high dose exposure pathway may be seasonal in this endemic population. The temporal clustering of symptomatic disease may further be a result of seasonal phage predation ^37^, serotype switching ^38^, or changing routes and/or manners of bacterial ingestion ^39^. Future detailed models exploring the forces behind these apparent different drivers of infection could ultimately improve our understanding of *V. cholerae* O1 dynamics.

Our results not only have implications for our understanding of the dynamics of this ancient disease but may provide important practical insights for the global fight against cholera and the End Cholera 2030 roadmap. Several countries have devised national cholera control plans that rely on passive clinical surveillance and have bold targets, often including elimination of the disease by 2030. Our results confirm previous modeling results that inapparent infections are key to understanding *V. cholerae* O1 transmission in endemic areas ^30^. This implies that the absence of cases may not mean absence of transmission, which must be taken into account when approaching elimination. After 3 years of no confirmed cases in Haiti, a new outbreak emerged in 2022 with genomic evidence linking the bacteria to the previously circulating 2010 strain in country, with cryptic transmission from subclinical infections (in the face of waning immunity and a lack of clean water sanitation) and an environmental reservoir being plausible explanations for this ^40^. Knowing the factors that shape this potential disconnect is likely critical for designing interventions for elimination, like vaccines and water and sanitation, assessing risk and establishing appropriate surveillance systems.

Though the most widely used cholera vaccines (killed whole oral vaccines) protect against severe disease, it is unclear how or if they protect against subclinical infection, particularly those that may lead to onward transmission ^41^. Defining the immunological markers of subclinical infections, correlates of protection, and the resulting different endpoints for clinical studies, can help us further understand the mechanisms of current vaccines and lead to the development of better ones in the years to come. Further, when exposures and risk are this pervasive, our results also strengthen the argument for safe, well-managed water systems. Individuals in our serologic cohort reported using piped water more often during the high transmission season compared to other times (Supplement Table S3), suggestive of a lack of consistent service and/or contamination ^42,43^. Though few cholera studies have estimated such a high proportion of subclinical infections, a comparably high infection to medically-attended case ratio (rate ratio of 16-32) has been documented for other enteric pathogens, like typhoidal *Salmonellae*, among Bangladeshi children ^44^ These concordant results could provide the impetus for aligned, cross-pathogen horizontal intervention mechanisms, specifically for clean water and sanitation.

Our study comes with several limitations. First, we based our model on the most influential marker of recent infection to estimate seroincidence, vibriocidal titers, though other antibody responses may contain additional information to help improve seroincidence estimates ^45^. We focused on vibriocidal titers given the existence of a well-characterized post-infection kinetic model from medically attended cholera cases calibrated on mostly adults ^22^. Future efforts may attempt to model the simultaneous decay of multiple markers ideally based on validation sets that include both mild and severe cholera infections. Second, our representative health seeking behavior survey results suggested that roughly one in four people with symptomatic cholera would seek care at one of the primary surveillance sites in Sitakunda for cholera like symptoms. This was based on the assumption that responses related to hypothetical moderate diarrhea represented the ‘average’ severity of disease, however, estimates of symptomatic incidence rates did vary (0.5-1.5 per 1,000 per year) when care-seeking was based on alternative diarrhea severity definitions (Supplement Table S4). It is likely that the clinical spectrum of disease varies across age groups (e.g., elderly and children more likely to have severe disease) and defining this distribution can aid in future work where these assumptions are needed.

Furthermore, the hypothetical care seeking questions we used in this survey may not capture the true behaviors of participants, likely overestimating people’s propensity to seek care. Larger studies asking about recent diarrheal events and subsequent care seeking could help improve our understanding of care seeking for cholera. Third, our inference in this study is based on data gathered from one year, 2021-2022, and in a single epidemiologic setting, therefore, the generalizability of these results temporally and geographically remains unclear.

Through combining serologic, epidemiologic, and behavioral data we decomposed the continuum between *V. cholerae* O1 infections and medically attended cholera cases observed within a typical surveillance system in an endemic community in Bangladesh. We reveal far higher than previously described rates of infection incidence with more than half the population infected each year and 3,020 infections per observed culture positive medically attended cholera case. The methods we developed here allowed us to map out a larger framework of burden estimation and scale the number of infection events in the community to symptomatic cases, medically attended cases, and test positive medically-attended cases that are ultimately counted within surveillance networks. These data support the relevance of quantifying subclinical infections in endemic settings so that we can better understand the natural history of *V. cholerae* O1 infection and adequately develop and allocate resources at the population-level to eliminate disease. Gaining a deeper understanding of subclinical infections in Bangladesh, where six of the last seven pandemics began, may be key to making serious progress in the global targets for cholera control.

## Methods

### Clinical and Laboratory Surveillance of Suspected Cholera Cases

Our study took place in the Sitakunda subdistrict (population size ∼441,676 for those ≥1 years) located in the Chattogram district of Bangladesh. Two public health facilities provide care for diarrheal patients: the Sitakunda sub-district hospital (Upazila Health Complex) and the Bangladesh Institute for Tropical Infectious Diseases (BITID). These two sites are the primary sites for surveillance of diarrheal disease and cholera for the Bangladesh Directorate General for Health Services in Sitakunda. From January 24, 2021 through February 13, 2022, we surveilled the in-patient and out-patient wards of both health facilities, and attempted to enroll all suspected cases ≥1 years old presenting with non-bloody, acute watery diarrhea (3 or more loose stools in the 24-hours preceding the visit). After obtaining informed consent we administered a short, structured questionnaire and collected a stool (or rectal swab) specimen for laboratory analyses. We tested each patient’s fecal sample onsite with the CholKit Rapid Diagnostic Test (RDT; Incepta, Dhaka, Bangladesh), then placed the sample in Cary Blair media and on Whatman 903 filter paper for subsequent lab testing at icddr,b in Dhaka. Due to the expected high sensitivity and moderate specificity of the RDT ^46^, all RDT positive samples were tested by culture and end-point PCR (for ctxA and rfb genes from filter paper) ^47^. Using the same PCR protocol, we additionally tested a random subset of nearly half of the RDT negative samples (46%). All diagnostic test results were used in a latent class model (*see* Supplement S4.1) to determine the joint weekly number of confirmed cases.

### Serologic Cohort and Care Seeking Survey

We enrolled a population representative cohort in Sitakunda between 27-March-2021 and 13-June- 2021, including a ∼1-month gap due to a national COVID-19 related lockdown (referred to as rounds R0A & R0B throughout). To enroll households, we used two-stage sampling based on satellite imagery (Airbus, Pléiades 1B sensor) with digitized building footprints classified as single or multi-story units, where we first divided the Sitakunda subdistrict into 1km^2^ grid-cells and then randomly selected grid-cells proportional to the number of households in each with replacement. Within each selected grid-cell, we randomly selected structures (or GPS coordinates) weighted by whether they were classified as single- or multi-story units; the number of structures selected by grid-cell varied and only one structure was enrolled per GPS coordinate selected. If no structure was found at the point or the structure located at the point was not residential, the study team attempted to enroll the nearest residential household within 20 meters or proceeded to the next assigned point if no residential household existed within 20 meters. If the structure found was multi-story, the study team enumerated the number of residential households inside and generated a random number to determine which household to attempt to enroll. If no one was at the household once it was found, the study team attempted to revisit up to three times in the following 24 hours. If no one was at home after the attempted revisits or the household refused to participate, the study team proceeded to the next point (or attempted to enroll the next closest residence to the right if in a multi-story unit).

After receiving verbal consent from the head of household (or representative), we attempted to enroll all persons ≥1 year of age that were members of the household (i.e., those who regularly sleep in the household and eat there) and asked for written consent (and assent for those 7-17 years old) from each person. When household members that met this inclusion criteria were not present, we attempted to revisit up to three times in the survey period to enroll them. Survey staff administered a structured household-level questionnaire to the head (or representative) of each enrolled household. They also administered an individual-level questionnaire to and collected venous blood from each consenting participant (∼5ml from adults and ∼3ml from children <5 years old). The baseline individual-level questionnaire included questions on each persons’ demographics, mobility patterns, health status and healthcare seeking behaviors. Each enrolled household had two follow-up visits at approximately 4-month intervals (R1 from 21-September-2021 to 9-October-2021 and R2 from 25- January-2022 to 13-February-20221). At each follow-up visit, members of households who were not previously enrolled (or declined to participate in the previous round) were eligible to participate, and all consenting participants were administered a follow-up individual-level questionnaire, venous blood draw, and household heads a follow-up household-level questionnaire. Follow-up questionnaires were aimed at recent clinical disease episodes, healthcare seeking and WASH-related behaviors like individual drinking water sources and household sanitation and water infrastructure.

To gauge people’s propensity to seek care for cholera, we asked a series of questions at baseline to each participant about three levels of hypothetical diarrhea severity: mild, moderate, and severe. We asked whether individuals would seek healthcare for each syndrome, and if yes, the specific type of care or facility (public, private, pharmacy, traditional healer, etc.) they would first visit. While the clinical spectrum of cholera is variable, we focused our main analyses on the self-reported care seeking for moderate-to-severe diarrhea (experiencing 3 or more loose stools in a day and dehydration) while exploring the other severities in sensitivity analyses.

Blood samples were centrifuged the day of collection and frozen at -80C until the time of analysis (1-11 months). Following previously described methods ^21^, we tested each sample with the vibriocidal assay, with all samples from the same participant run on the same plate. Serum samples collected at different rounds were tested for vibriocidal antibody response against the homologous serotype of bacteria using the *V. cholerae* O1 El Tor Ogawa (strain 25049) or Inaba (strain 19479) strains as the target organism ^48^. Guinea pig complement (Sigma, USA) was used at a 1:10 dilution. The ODs of the plates were measured at 595 nm. A known pooled sera (prepared from convalescent sera of 20-30 cholera patients) was used in each plate as a control. The vibriocidal titer was defined as the reciprocal of the highest serum dilutions causing a greater than 50% reduction of the OD at 595 nm when compared with the OD of the control wells without serum. Wells containing physiological saline, and growth medium were included on each plate to exclude the possibility of bacterial contamination of the reagents.

### Sitakunda Population Estimates

We used the reported population size by 5-year age bins for Sitakunda from the 2011 National Census^49^. We extrapolated population counts by age group and sex assuming 1.5% annual population growth for the 10 years between 2011 and 2021. Since children under one year of age were excluded in the survey and clinical surveillance, we subtracted 20% of the total population for the 0-4 age group, following the age distribution reported in the US Census Bureau International Database for Bangladesh in 2021 ^50^.

### Reconstruction of Medically Attended Cholera Incidence Rates

We estimated the age-stratified incidence rates of medically attended cholera and non-cholera suspected cases within our study facilities using a Bayesian model fit to data from systematic laboratory testing of all suspected cases. Data were restricted to individuals residing in Sitakunda subdistrict visiting the healthcare facilities from the start of the study period until February 13, 2022, the end of the last serosurvey. In short, we assume that the incidence rate of acute water diarrhea (*β_awd_*) can be decomposed into the sum of the incidence rates of cholera (*β_chol_*) and non-cholera (*β*_∼*chol*_) AWD. We further assume that both the incidence rates of cholera and non-cholera AWD evolved over time during the study period following a first order Brownian motion process at the daily time-scale. We link the model to data by assuming that the total number of daily AWD cases follows a Poisson distribution with rate *β_awd_*. Given that we tested all suspected cases with RDTs, which have imperfect sensitivity and specificity ^46^, and in some cases PCR and/or culture, which are also imperfect ^51^, we modeled the array of test outcomes using a multinomial likelihood, marginalizing over potential outcomes of tests not performed while accounting for the conditional dependence between the tests inherent in study design (i.e., testing of PCR/culture based on RDT results) with priors on performance derived from a prior study in Bangladesh ^51^. The result of this model component is the posterior distribution of the age-group specific daily incidence rate of cholera and non-cholera AWD from study health facilities. Complete details of this model are provided in the supplement (Supplement S4).

### Seroincidence Estimation Framework

Our estimates of infection incidence are based on serial measurements of vibriocidal antibodies developed in response to *V. cholerae* O1 Ogawa, the predominant regional serotype ^52^, from each participant. While simple definitions of seroincidence typically include (a) any rise in vibriocidal titer between visits or (b) any rise greater than a threshold (e.g., 2-fold rise), these will be biased as titers decay quickly, often within months, and thus in a shorter time frame than our study visits. Also, these potential definitions do not account for measurement error in the vibriocidal assay. We, therefore, developed a Bayesian inference framework to estimate the seroincidence rate in the population based on serial measurements accounting for the decay kinetics of vibriocidal antibodies, the observed measurement error in the assay and the reconstructed epidemic curve of medically attended cholera cases as a proxy for the force of infection experienced by study participants. We restricted these analyses to only those who participated in all three study visits (72% of the total enrolled).

We assumed that post-infection antibody kinetics followed a previously published antibody kinetics model based on data from medically attended cholera patients in Dhaka, Bangladesh ^22^. In this model, vibriocidal antibodies were assumed to have an instantaneous boost after some delay post-infection from their baseline levels and then decay exponentially at a constant rate. The differences (or lack thereof) in vibriocidal measurement between rounds could indicate (1) absence of recent infection with any differences due to measurement error, (2) prior infection (before the last study visit) leading to a decay in vibriocidal titers, or (3) an infection at some unknown time since the last visit potentially with some decay in titer levels. Our inference framework estimates the likelihood of each of these possibilities for each person-sample, while utilizing the data from clinical surveillance to infer the likelihood of infections at each timepoint between measurements.

The likelihood function is based on eight potential outcomes for individual-level infection status before baseline and between each study round for each participant (e.g., <0,0,0> indicates that the participant was not ‘recently’ infected before any of the study visits and <0,1,0> indicates that the participant was infected between the baseline and first visits). We can then estimate the probability of infection between any two arbitrary time points (*t_a_* and *t_b_*) for a person, *i*, as a function of the force of infection, which we assume the time-varying incidence rate of medically attended cholera (*β_chol_*) is a valid proxy of, scaled by a constant factor (*λ*) as follows:

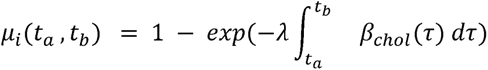

In this formulation, we implicitly assume that infections in the community overall are a function of the symptomatic cholera incidence leading to observed medically attended cases in the clinics. We also considered a second model where seroincidence is a function of two components, the force of infection proportional to inferred symptomatic community infections and a constant force of infection independent of cases in the clinics:

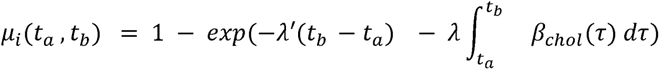

We can link cumulative medically attended cholera cases to cumulative seroincidence through the combined probabilities of symptomatic infection given infection, *ϕ*, and the probability of health seeking given symptomatic infection (*δ_a_*) as:

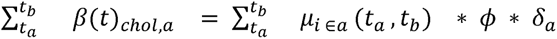

We infer cumulative cases in the first inference step (Supplement Figure S4), cumulative seroincidence in the second, and estimate posterior estimates from our health seeking behavior survey; thus, the remaining unknown is the probability of symptomatic infection, which we estimate here (Supplement Figure S5).

We fit these models using a Hamiltonian Markov chain algorithm as implemented in Stan and assessed model convergence through visual inspection of trace plots and use of the Rhat statistics. We performed several posterior predictive checks to understand model fit and compared model formulations using an estimate of leave-one-out cross-validation performance (LOO-PSIS) ^53^. Complete details are provided in the supplement.

## Data Availability

All data produced in the present study are available upon reasonable request to the authors.

## Supplementary Figures and Tables

*See manuscript/supplement/chol_incidence_supplement.pdf doc for materials*.

## Funding and Acknowledgements

We thank all the participants in this study and our collaborators in the Sitakunda sub-district for making this work possible. This work was supported by a grant from the Bill and Melinda Gates Foundation (INV-021879). Andrew Azman received support from the US National Institutes of Health (R01-AI135115). The icddr,b is grateful to the governments of Bangladesh, and Canada for providing core/unrestricted support.

## Notes

### Competing Interest Statement

The authors have declared no competing interest.

### Author Declarations

Our study was approved by the icddr,b research and ethics review committee and the Johns Hopkins Bloomberg School of Public Health institutional review board.

