## Supplemental material for "Estimating the gap between clinical cholera and true community infections: findings from an integrated surveillance study in an endemic region of Bangladesh"

### Supplementary material

#### Contents

**S1 Survey description** **S2**

**S2 Health seeking statistics** **S4**

**S3 Vibriocidal titers** **S5**

**S4 Modeling framework** **S6**

S4.1 Time varying force of infection and weekly cholera incidence rates . . . . . . . . . . . . . . . . S7

S4.2 Titer trajectories . . . . . . . . . . . . . . . . . . . . . . . . . . . . . . . . . . . . . . . . . . . . . . . S12

**S5 Medically-attended suspected case to true cholera case ratio** **S16**

**S6 Drinking water sources** **S17**

**S7 True symptomatic *V. cholerae* incidence for different definitions of diarrhea severity** **S17**

### S1 Survey description

This study was conducted in the Sitakunda subdistrict of the Chattogram district of Bangladesh. We enrolled a population representative cohort from 580 households across the subdistrict. We also conducted clinical surveillance at the two primary healthcare facilities in the subdistrict where patients with cholera-like symptoms would visit. The Sitakunda Upazila Health Complex (Sitakunda UHC) is located towards the north of the subdistrict and the Bangladesh Institute of of Tropical and Infectious Diseases (BITID) is located towards the southern end of the subdistrict, near the city of Chittagong. Among the 2,176 suspected cases that visited the study clinical facilities during the study period, 99% (N=2,158) were from or spent the last 7 days in the Chattogram district. Among the suspected cases from the Chattogram district (N=2,158), 46% (N=998) were from or spent the last 7 days in the subdistrict of Sitakunda.

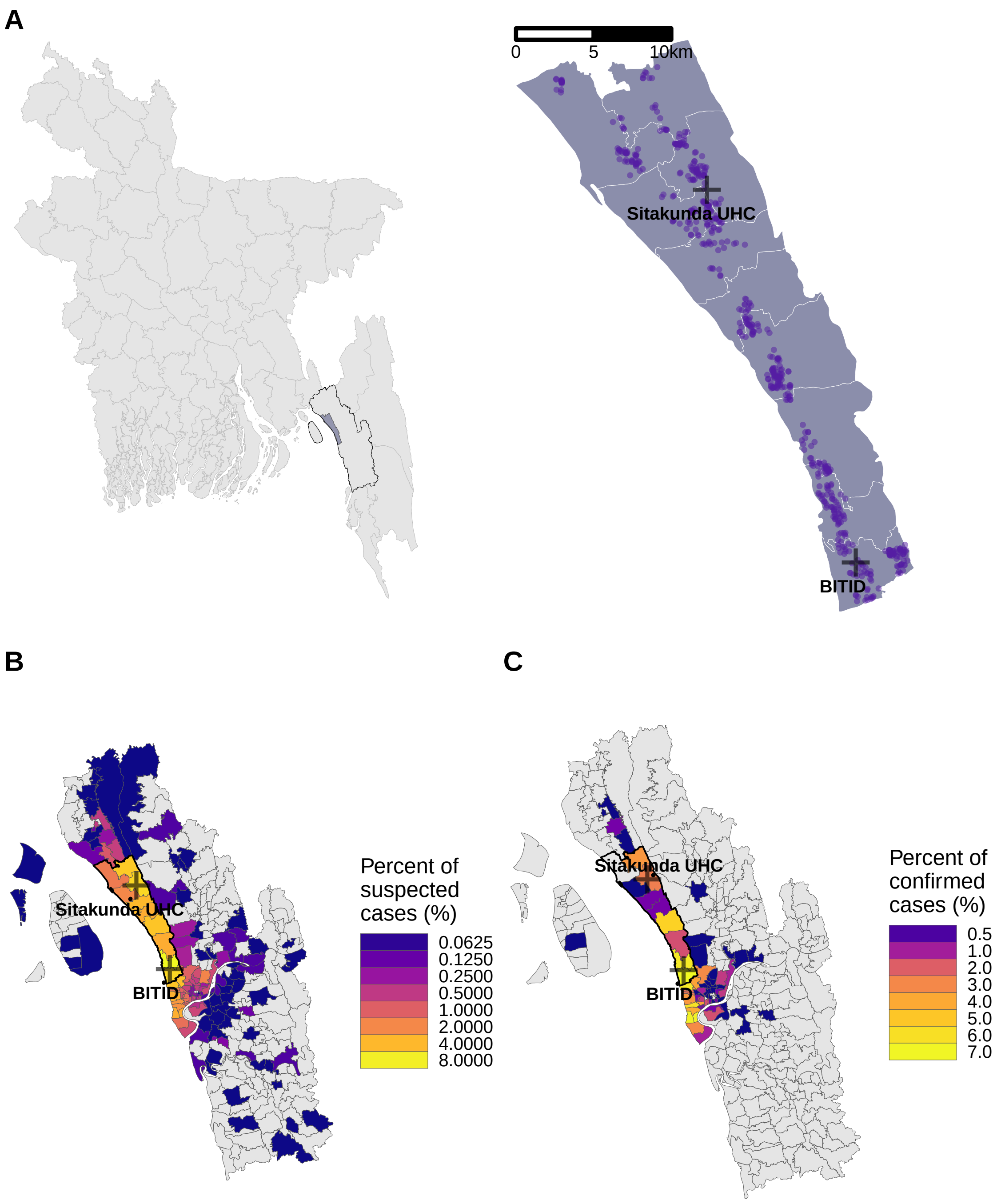


Figure S1: Map of the enrolled households in the serosurvey in the Sitakunda sub-district (A), and the suspected (B) and confirmed (C) clinical V. cholerae cases by RDT test result by Union (the lowest administrative unit) in the Chattogram district of Bangladesh.

We enrolled 2,481 participants into the baseline survey, and followed all that consented (N=1,785) as a serological cohort to capture incident serologically-defined infections. The age and sex profile of the enrolled serological cohort matched that of the national population with notable undersampling of young children.

The enrolled participants in clinical surveillance, both those who were enrolled as suspected cholera cases and confirmed cholera positive by RDT, roughly followed the age and sex distribution of the national population. However, children less than five years old were overrepresented when compared to the population and those 5-19 years old were under represented.

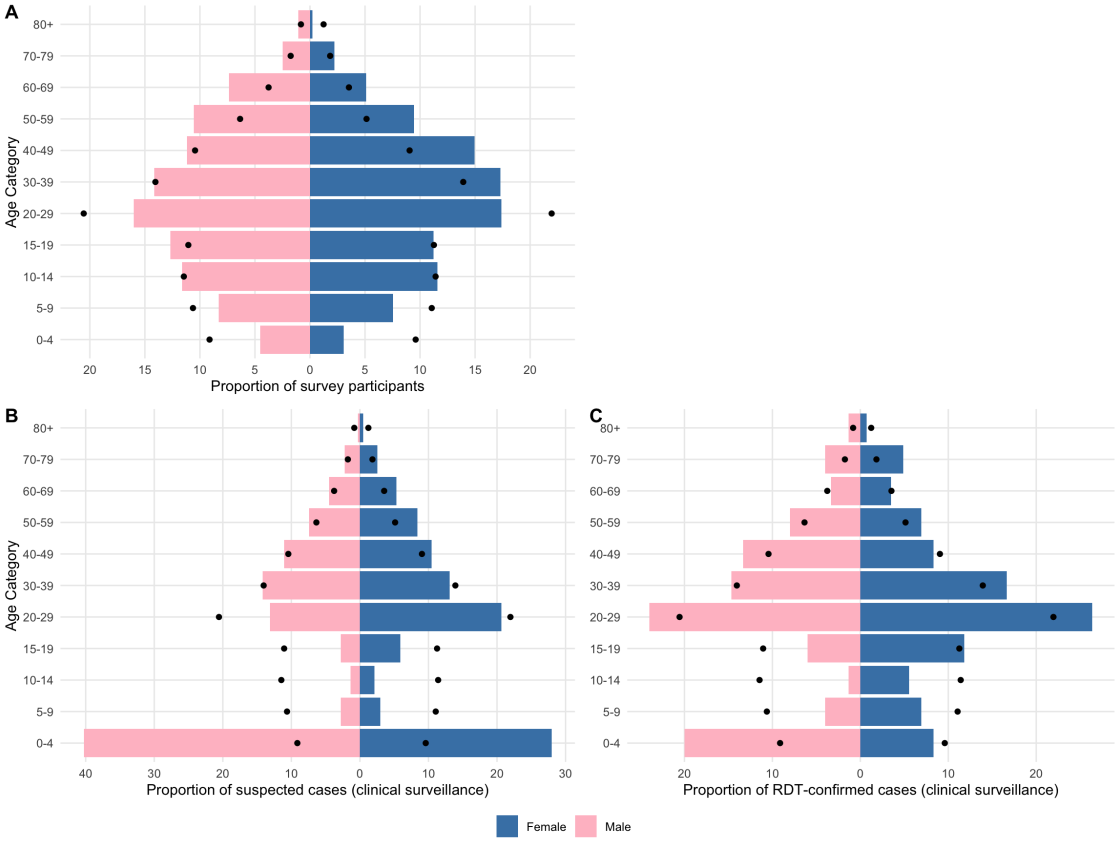


Figure S2: Demographic pyramid of survey participants (only those with three follow-up visits) and of confirmed and suspected cases.

#### S2 Health seeking statistics

We asked hypothetical healthcare seeking questions to all baseline participants for different levels of diarrheal disease severity. To gauge healthcare seeking propensity for mild, moderate, and severe diarrhea, participants were asked the following questions:

- Mild diarrhea: After experiencing **3 or more loose stools in 1 day**, would you seek medical care?
- Moderate diarrhea: After experiencing **3 or more loose stools in a day and dehydration**, would you seek medical care?
- Severe diarrhea: After experiencing **3 or more loose stools per day for more than 3 days**, would you seek medical care?

Participants were also asked which facility they would visit if they would seek care for each syndrome, including if they would seek care at the two health facilities under surveillance in the study. There were no significant differences in healthcare seeking for moderate diarrhea; 26.1% (n=29) of participants 1-4 years of age, 25.9% (n=576) of participants 5-64 year of age, and 24.3% (n=36) of participants older than 65 years said they would seek care at either Sitakunda UHC or BITID (Table S1).

Table S1: Proportion of survey particpants reporting that they would seek care for different diarrhea severities at one of the two official diarrhea treatment centres in the catchment area (Sitakunda Upazila Health Complex or BITID).

| Age group | N | Seek health='yes' | Proportion [mean (95% CrI)] |
|---|---|---|---|
| **Mild diarrhea** | | | |
| $< 5$ | 111 | 22 | 0.20 (0.13-0.28) |
| $5 - 64$ | 2221 | 330 | 0.15 (0.13-0.16) |
| 65+ | 148 | 16 | 0.11 (0.07-0.17) |
| **Moderate diarrhea** | | | |
| $< 5$ | 111 | 29 | 0.27 (0.19-0.35) |
| $5 - 64$ | 2222 | 576 | 0.26 (0.24-0.28) |
| 65+ | 148 | 36 | 0.25 (0.18-0.32) |
| **Severe diarrhea** | | | |
| $< 5$ | 111 | 54 | 0.49 (0.40-0.58) |
| $5 - 64$ | 2222 | 966 | 0.43 (0.41-0.46) |
| 65+ | 148 | 63 | 0.43 (0.35-0.51) |

### S3 Vibriocidal titers

The serologic cohort data provided serum samples from three different time-points per individual, and these serum samples were tested using the vibriocidal assay to track changes in individuals' antibodies against *Vibrio cholerae* O1 over the study period. The overall distribution of vibriocidal antibodies was similar across rounds of data collection, with 18.7% of participants having a titer greater than or equal to 320 at baseline (during both enrollment periods; 17.6% during R0A and 19.1% during R0B), 21% during the first follow-up (R1), and 19.4% during the second follow-up (R2). Overall, 8.1% of participants had a greater than or equal to 2-fold rise (defined as log2 difference in titers greater than or equal to 2) in vibriocidal titers across any two study visits including 14.9% among children 1-4 years old at baseline, 7.8% among those 5-64 years old and 8.6% among those 65 years and older. Overall, 4% of individuals had a greater than or equal to 2-fold rise in vibriocidal from enrollment to their first follow-up visit, and 4.2% had a greater. than or equal to 2-fold rise between the first follow-up and the final visit (Figure S3).

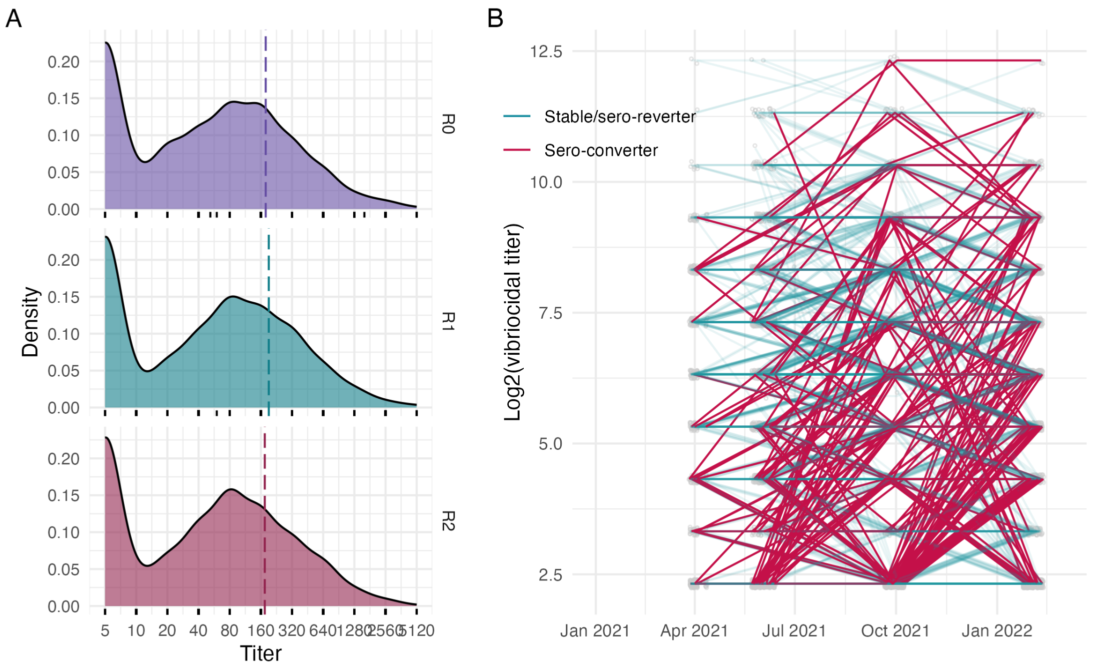


Figure S3: A. Density plot of Vibrio cholerae O1 Ogawa vibriocidal titers by serosurvey round where the dashed line indicates the population mean titer for the N=1,785 individuals that provided serum samples for all three rounds of data collection. The rug plots below indicate the actual titer values measured. B. The trajectory of log2(vibriocidal titers) across serosurvey rounds grouped by those who sero-convert (greater than or equal to 2 fold rise in titer value across any two study visits) and those who remain stable or sero-revert.

### S4 Modeling framework

Here we provide details on our framework for estimating the cascade from total *V. cholerae* O1 infections in the community, to resulting symptomatic infections and reported clinical cholera cases (Figure S4). The analysis relies on the inference of seroincidence rates based on serial measurements of vibriocidal antibodies. In turn, this inference relies on estimates of the time-varying force of infection from *V. cholerae* O1, which results in seroconversions in between the study rounds.

The modeling framework consists of two primary parts (1) reconstruction of the time-varying force of infection and weekly medically-attended cholera incidence rates from the clinical surveillance data and (2) modeling of probable titer trajectories in between study visits accounting for infection-induced antibody boosts conditional on a known antibody kinetic model. These two parts are connected through the force of infection of *V. cholerae* infections, which influences the probability of seroconversion between blood draws (i.e., study rounds) for each participant (Figure S4).

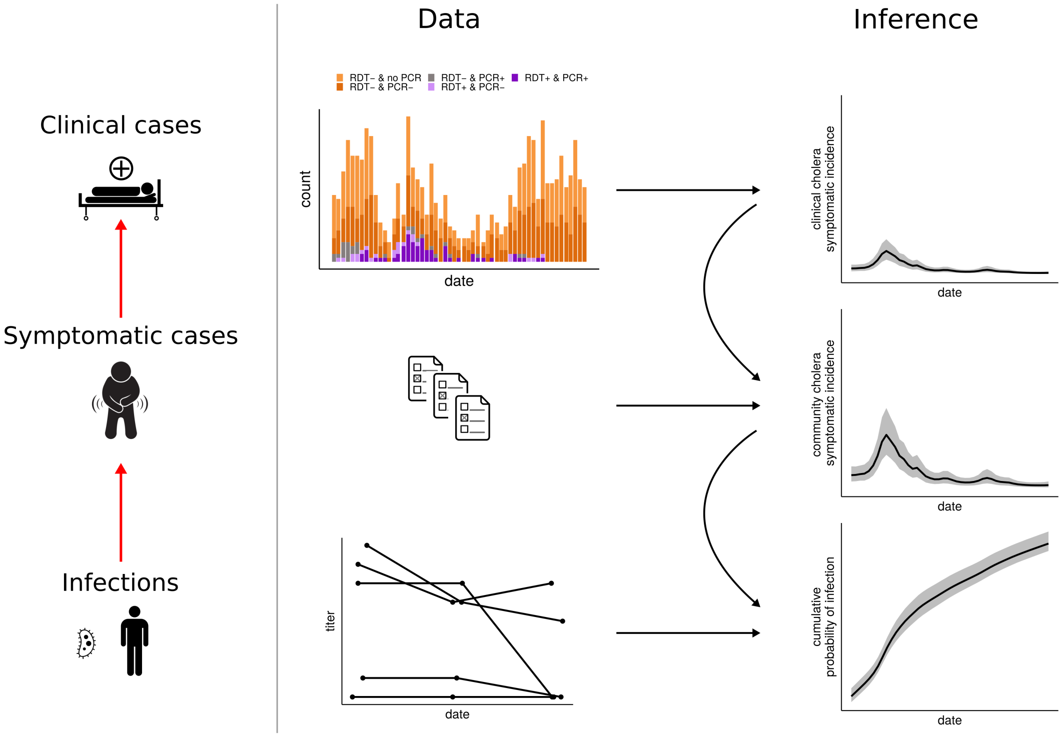


Figure S4: Study aim and modeling framework.

#### S4.1 Time varying force of infection and weekly cholera incidence rates

##### S4.1.1 Base model

To model the time-varying force of infection of cholera we use clinical surveillance data of suspected and confirmed cholera cases. Let $\beta_{chol}(t)$ and $\beta_{\neg chol}(t)$ be the daily incidence rates of cholera and non-cholera acute watery diarrhea (AWD) recorded in the health centers. Both are time-varying, and are assumed to follow a first-order Brownian motion with initial value $\alpha$:

$$\begin{aligned}
\beta_{chol}(t) &= \exp(\alpha_{chol} + \Delta_t \log(\beta_{chol})), \\
\Delta_t \log(\beta_{chol}) &= \log(\beta_{chol}(t+1)) - \log(\beta_{chol}(t)) \sim \mathcal{N}(0, \sigma_{\beta,chol}), \\
\beta_{\neg chol}(t) &= \exp(\alpha_{\neg chol} + \Delta_t \log(\beta_{\neg chol})), \\
\Delta_t \log(\beta_{\neg chol}) &= \log(\beta_{\neg chol}(t+1)) - \log(\beta_{\neg chol}(t)) \sim \mathcal{N}(0, \sigma_{\beta,\neg chol}), \\
\beta_{awd}(t) &= \beta_{chol}(t) + \beta_{\neg chol}(t), \\
\phi(t) &= \frac{\beta_{chol}(t)}{\beta_{awd}(t)}, \\
n_{awd}(t) &\sim Poisson(\beta_{awd}(t)),
\end{aligned}$$

where $\beta_{awd}$ is the total AWD daily incidence, and $\phi$ is the time-varying proportion of AWD incidence due to cholera. The number of AWD cases is assumed to follow a Poisson distribution with rate $\beta_{awd}$. The likelihood is complemented by the probability of observing specific cholera test results, as described in the next section.

##### S4.1.2 Accounting for multiple tests

The surveillance scheme implemented in the study is described in the main text. Briefly, all suspected cholera cases were tested with RDT, and a subset was also tested with PCR and culture. We first assume that all three test results are available for all samples, and then extend to the case where they are not. To account for different potential outcomes we model the joint weekly result of RDT, PCR and culture as a multinomial distribution:

$$\begin{aligned}&\left[n_{\{-,-,-\}}, n_{\{-,-,+\}}, n_{\{-,+,-\}}, n_{\{-,+,+\}}, n_{\{+,-,-\}}, n_{\{+,-,+\}}, n_{\{+,+,-\}}, n_{\{+,+,+\}}\right] \sim \\ &multinomial(p_{\{-,-,-\}}, p_{\{-,-,+\}}, p_{\{-,+,-\}}, p_{\{-,+,+\}}, p_{\{+,-,-\}}, p_{\{+,-,+\}}, p_{\{+,+,-\}}, p_{\{+,+,+\}}),\end{aligned}$$

where signs in brackets indicate the result of RDT, PCR and culture respectively (e.g., $\{-,+,-\}$ indicates a negative RDT, a positive PCR and a negative culture result).

Each test tupple of outcome probabilities are a function of the true probability of cholera $\phi$ among suspected cases (i.e. the fraction of total AWD due to cholera), and the test performances. Assuming that the different test outcomes of a suspected cholera case's are independent conditional on the probability of cholera, and that all test outcomes are known, the corresponding probabilities are:

$$\begin{aligned}
p_{\{-,-,-\}} =&(1-\theta_1^+)(1-\theta_2^+)(1-\theta_3^+)\phi + \theta_1^-\theta_2^-\theta_3^-(1-\phi),\\
p_{\{-,-,+\}} =&(1-\theta_1^+)(1-\theta_2^+)\theta_3^+\phi + \theta_1^-\theta_2^-(1-\theta_3^-)(1-\phi),\\
p_{\{-,+,-\}} =&(1-\theta_1^+)\theta_2^+(1-\theta_3^+)\phi + \theta_1^-(1-\theta_2^-)\theta_3^-(1-\phi),\\
p_{\{-,+,+\}} =&(1-\theta_1^+)\theta_2^+\theta_3^+\phi + \theta^-(1-\theta_2^-)(1-\theta_3^-)(1-\phi),\\
p_{\{+,-,-\}} =&\theta_1^+(1-\theta_2^+)(1-\theta_3^+)\phi + (1-\theta_1^-)\theta_2^-\theta_3^-(1-\phi),\\
p_{\{+,-,+\}} =&\theta_1^+(1-\theta_2^+)\theta_3^+\phi + (1-\theta_1^-)\theta_2^-(1-\theta_3^-)(1-\phi),\\
p_{\{+,+,-\}} =&\theta_1^+\theta_2^+(1-\theta_3^+)\phi + (1-\theta_1^-)(1-\theta_2^-)\theta_3^-(1-\phi),\\
p_{\{+,+,+\}} =&\theta_1^+\theta_2^+\theta_3^+\phi + (1-\theta_1^-)(1-\theta_2^-)(1-\theta_3^-)(1-\phi),
\end{aligned}$$

where $\theta^+$ and $\theta^-$ are the test sensitivity and specificity, and subscripts 1,2,3 denote RDT, PCR and culture, respectively.

##### S4.1.3 Accounting for partial testing

If data were available for all tests and samples then the likelihood of the sequence of samples could be expressed as a product of multinomial probabilities as described above. However our sampling protocol consisted of partial PCR and culture testing as a function of the initial RDT result. Specifically, results could be in one of three cases: i) only RTD- results (four in five RDT- tests), ii) RDT- result plus PCR test result (one in five RDT- result), and iii) RDT, PCR and culture result (all RDT+ tests).

If partial testing had been completely at random, it would be possible to directly integrate out missing test results assuming a uniform prior over missing test outcomes. Given that our sampling scheme depended on the initial RDT result the uniform prior assumption does not hold and we need to account for the prior probability of test results conditional on the initial test result. Case (iii) (all tests were performed) does not require marginalization since all three test were performed, and the observation likelihood can be directly obtained through the multinomial likelihood detailed above. Cases (i) and (ii) on the other hand require marginalization.

**Case (i) (only RDT- result available)**: Let $Y_1, Y_2, Y_3 \in \{0, 1\}$ denote the binary random variables representing the results of RDT, PCR and culture respectively. The prior probability of a given PCR, $Y_2 = y_2$, and culture, $Y_3 = y_3$, test result given a negative RDT result, $Y_1 = 0$, is:

$$Pr(Y_2 = y_2, Y_3 = y_3 | Y_1 = 0) = \frac{Pr(Y_1 = 0, Y_2 = y_2, Y_3 = y_3)}{Pr(Y_1 = 0)}.$$

We can obtain the numerator and denominators of the right hand side of this equation above by integrating out the unobserved cholera infection status, $x \in \{0, 1\}$ as:

$$\begin{aligned} Pr(Y_1 = 0, Y_2 = y_2, Y_3 = y_3) &= \sum_{x\in\{0,1\}} Pr(Y_1 = 0, Y_2 = y_2, Y_3 = y_3 | x) Pr(x) \\ &= \sum_{x\in\{0,1\}} Pr(Y_1 = 0 | Y_2 = y_2, Y_3 = y_3, x) Pr(Y_2 = y_2 | Y_3 = y_3, x) Pr(Y_3 = y_3 | x) Pr(x), \\ &= \sum_{x\in\{0,1\}} Pr(Y_1 = 0 | x) Pr(Y_2 = y_2 | x) Pr(Y_3 = y_3 | x) Pr(x) \end{aligned}$$

where $Pr(x)$ is the prior probability of cholera infection. The last equality is obtained by assuming that test results for a given sample are independent conditional on the participant's infection status.

We can then use the above to complete the probability of the unobserved tests given a negative RDT test as follows:

$$\begin{aligned} Pr(Y_2 = y_2, Y_3 = y_3 | Y_1 = 0) &= \frac{Pr(Y_1 = 0, Y_2 = y_2, Y_3 = y_3)}{Pr(Y_1 = 0)} \\ &= \frac{\sum_{x\in\{0,1\}} Pr(Y_1 = 0|x) Pr(Y_2 = y_2|x) Pr(Y_3 = y_3|x) Pr(x)}{\sum_{x\in\{0,1\}} Pr(Y_1 = 0|x) Pr(x)} \\ &= \Phi_{\{-,y_2,y_3\}} \end{aligned}$$

where we use $\Phi_{\{-,y_2,y_3\}}$ to denote the prior probability of PCR and culture results $y_2$ and $y_3$ conditional on the negative RDT test result.

The probabilities in the equation above can then be expressed in terms of the test sensitivity, $\theta^+_{1,2,3}$ and specificity $\theta^-_{1,2,3}$ as:

$$\begin{aligned} &Pr(Y_i = 0 | x = 0) = \theta_i^-, \\ &Pr(Y_i = 1 | x = 0) = 1 - \theta_i^-, \\ &Pr(Y_i = 0 | x = 1) = 1 - \theta_i^+, \\ &Pr(Y_i = 1 | x = 1) = \theta_i^+. \end{aligned}$$

Priors on test performance were taken from an evaluation of the same RDT, Cholkit, used in this study conducted in Dhaka Bangladesh (Sayeed et al. (2018)).

Finally the likelihood of an RDT- result in case (i) given the probability vector $p$ can be expressed by marginalizing out the PCR and culture results accounting for their prior probabilities of occurrence:

$$Pr(\{-, \cdot, \cdot\} | p) = \sum_{y_2\in\{0,1\}, y_3\in\{0,1\}} multinomial(\boldsymbol{n}_{\{-,y_2,y_3\}} | p) \Phi_{\{-,y_2,y_3\}},$$

were $\boldsymbol{n}_{\{-,y_2,y_3\}}$ is the corresponding indicator vector for the outcome of interest.

**Case (ii) (RDT- and PCR result available)**: We can derive this case in a similar manner by considering the conditional probability for a given known PCR test result, and marginalizing out the unobserved culture result. Specifically:

$$
\begin{aligned}
Pr(Y_3 = y_3 | Y_1 = 0, Y_2 = y_2) &= \frac{Pr(Y_1 = 0, Y_2 = y_2, Y_3 = y_3)}{Pr(Y_1 = 0, Y_2 = y2)} \\
&= \frac{\sum_{x\in\{0,1\}} Pr(Y_1 = 0|x) Pr(Y_2 = y_2|x) Pr(Y_3 = y_3|x) Pr(x)}{\sum_{x\in\{0,1\}} Pr(Y_1 = 0|x) Pr(Y_2 = y_2|x) Pr(x)} \\
&= \Phi'_{\{-,y_2,y_3\}}.
\end{aligned}
$$

We then have for case (ii):

$$
Pr(\{-, y_2, \cdot\}|p) = \sum_{y_3\in\{0,1\}} multinomial(\boldsymbol{n}_{\{-,y_2,y_3\}}|p)\Phi'_{\{-,y_2,y_3\}}.
$$

In the implementation of these prior probabilities we set the prior probability of cholera to $Pr(x) = 0.2$.

###### S4.1.4 RDT batch effects

We separate the modeling period into two distinct periods, one from the start of the study up to June 29th 2021, and one from June 30th to the end of the study period.

###### S4.1.5 Age-specific model

In the model above we assume that all age classes experience the same incidence rate. Given observed differences in AWD cases and RDT positivity rates we choose to model age classes separately. We therefore subdivide clinical samples into three age groups: below 5, between 5 and 64, and 65 years and above. We then expand the time varying force of infection model to have age-specific cholera and non-cholera incidence. We assume that test performance is the same across age classes.

###### S4.1.6 Priors

We use the following priors in the cholera incidence model with no differences between age classes:

$$
\begin{aligned}
\alpha_{chol} &\sim \mathcal{N}(0, 1), \\
\alpha_{\neg chol} &\sim \mathcal{N}(0, 1), \\
\sigma_{chol} &\sim \mathcal{N}(0, 1), \\
\sigma_{\neg chol} &\sim \mathcal{N}(0, 1), \\
logit(\theta^{+}_{RTD_1}) &\sim \mathcal{N}(0, 0.75), \\
logit(\theta^{+}_{RTD_2}) &\sim \mathcal{N}(3.89, 0.95), \\
logit(\theta^{+}_{RT-PCR}) &\sim \mathcal{N}(1.04, 0.34), \\
logit(\theta^{+}_{culture}) &\sim \mathcal{N}(0.89, 0.26), \\
logit(\theta^{-}+_{RTD_1}) &\sim \mathcal{N}(3.48, 0.69), \\
logit(\theta^{-}_{RTD_2}) &\sim \mathcal{N}(3.48, 0.69), \\
logit(\theta^{-}_{RT-PCR}) &\sim \mathcal{N}(3.48, 0.44), \\
logit(\theta^{-}_{culture}) &\sim \mathcal{N}(6.9, 0.55).
\end{aligned}
$$

###### S4.1.7 Posterior retrodictive checks

We conducted retrodictive checks to understand how well the model could reproduce the testing data. The following plot illustrates the data (red) and the predicted values and 95% prediction intervals in black.

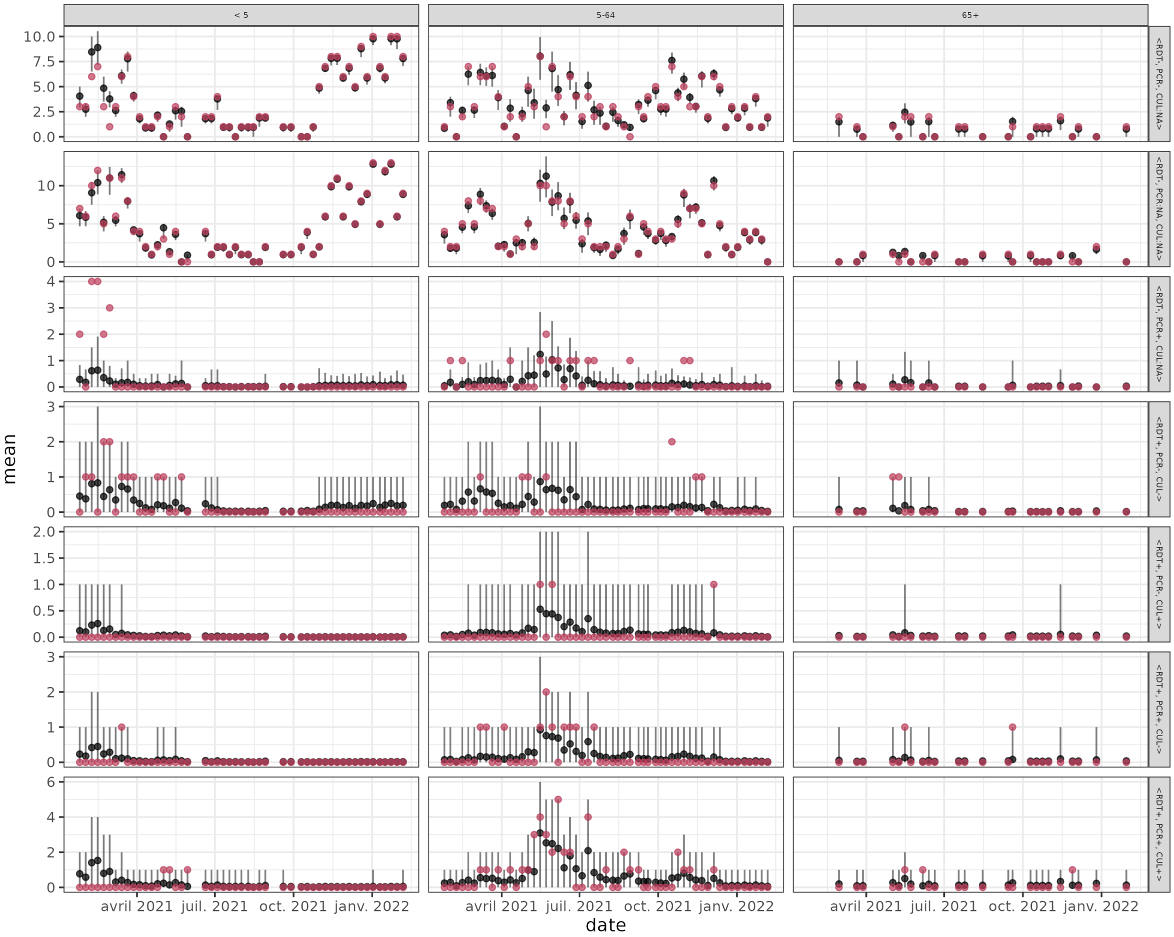


Figure S5: Posterior retrodictive checks of surveillance data.

##### S4.1.8 Key posterior parameter draws

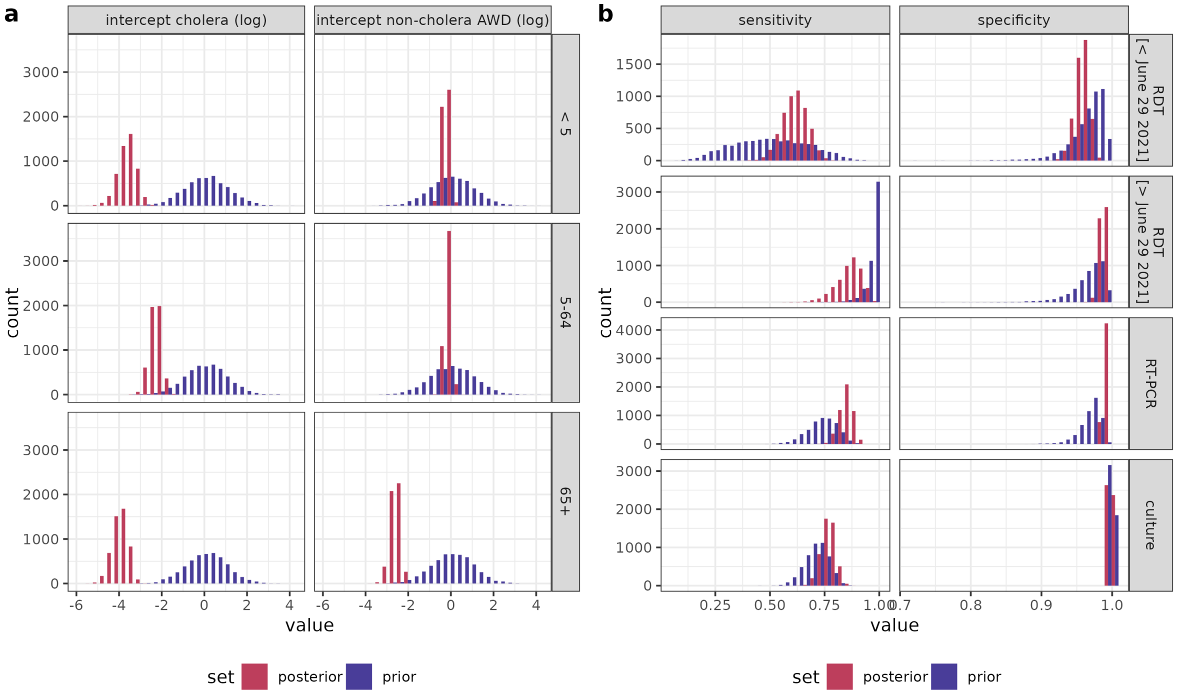


Figure S6: Clinical incidence model key parameter prior and posterior draws. a) Histograms of intercepts of cholera and non-cholera AWD daily incidence (on log scale). b) Histograms of test sensitivity and specificity.

#### S4.2 Titer trajectories

##### S4.2.1 Model description

Vibriocidal titer trajectories were assumed to follow exponential decay as estimated in Jones et al. (2022). To infer the probabilities of being infected at each serosurvey round, we marginalize over possible infection dates before baseline, and between each serosurvey round. The observation likelihood is then composed of the titer observation likelihoods assuming the decay model, weighted by the probability of being infected in each period.

For a participant $i$, let $\mu_{i,T}$ be the probability of infection in a given time period $T$, and $y_{i,r}$ denote a serological measurement made at serosurvey round $r$. In this study, we define three periods corresponding to the three serological measurement rounds. The first covers the six month preceding baseline (round 1), the second between baseline and round 2, and the third between round 2 and round 3. We denote by $\mu_{\{0,0\to1,1\to2\}}$ and $y_{\{1,2,3\}}$ the corresponding infection probabilities and serological measurements. We denote by $x(t,\tau)$ the modeled vibriocidal titer measured at time $t$ with exposure at a previous time $t-\tau$, following an exponential decay kinetic model as in Jones et al. (2022). Briefly, these models assume that antibody titers increase following exposure by a given boost value after a given delay, and then decay exponentially to a pre-exposure baseline level. Let $x_r^\star(t)$ denote the modeled vibriocidal titer assuming the baseline titer is equal to 0. Modeled antibody titer trajectories are linked to serological measurement through the probability density function $f_{Y|X}(y|x,\theta)$ with parameters $\theta$. We here choose this observation model to be a normal distribution (on the log scale of titers) with a known standard deviation $\sigma$ (based on the variability of positive controls across plates within experiments done for this study), which we denote as $f_N(y|x)$ in the following. We finally denote as $p_{t_a}^{t_b}(t)$ the prior conditional probability of infection at time $t_a \leq t \leq t_b$, given that infection did occur.

With these notations, we can then define the eight possible sequences of infection statuses during each period:

$$< 0,0,0 > : (1-\mu_0)(1-\mu_{0\to 1})(1-\mu_{1\to 2}) \times \prod_{r=1}^{3} f_N(y_{i,r}|\gamma),$$

$$< 0,0,1 > : (1-\mu_0)(1-\mu_{0\to 1})\mu_{1\to 2} \times \prod_{r=1}^{2} f_N(y_{i,r}|\gamma) \times \int_{t_2}^{t_3} f_N(y_{i,3}|x(t_3,\tau))p_{t_2}^{t_3}(\tau)d\tau,$$

$$< 0,1,0 > : (1-\mu_0)\mu_{0\to 1}(1-\mu_{1\to 2}) \times f_N(y_{i,1}|\gamma) \times \int_{t_1}^{t_2} f_N(y_{i,2}|x(t_2,\tau))f_N(y_{i,3}|x(t_3,\tau))p_{t_1}^{t_2}(\tau)d\tau,$$

$$< 0,1,1 > : (1-\mu_0)\mu_{0\to 1}\mu_{1\to 2} \times f_N(y_{i,1}|\gamma)\times$$
$$\int_{t_1}^{t_2}\int_{t_2}^{t_3} f_N(y_{i,2}|x(t_2,\tau))f_N(y_{i,3}|x(t_3,\tau)+x^\star(t_3,\tau'))p_{t_1}^{t_2}(\tau)p_{t_2}^{t_3}(\tau')d\tau d\tau',$$

$$< 1,0,0 > : \mu_0(1-\mu_{0\to 1})(1-\mu_{1\to 2}) \times \prod_{r=1}^{3}\int_{t_{r-1}}^{t_r} f_N(y_{i,r}|x(t_r,\tau))p_{t_{r-1}}^{t_r}(\tau)d\tau,$$

$$< 1,0,1 > : \mu_0(1-\mu_{0\to 1})\mu_{1\to 2}\prod_{r=1}^{2}\int_{t_0}^{t_1} f_N(y_{i,r}|x(t_r,\tau))p_{t_0}^{t_1}(\tau)d\tau\times$$
$$\int_{t_0}^{t_1}\int_{t_2}^{t_3} f_N(y_{i,2}|x(t_2,\tau))f_N(y_{i,3}|x(t_3,\tau)+x^\star(t_3,\tau'))p_{t_0}^{t_1}(\tau)p_{t_2}^{t_3}(\tau')d\tau d\tau',$$

$$< 1,1,0 > : \mu_0\mu_{0\to 1}(1-\mu_{1\to 2}) \times \int_{t_0}^{t_1} f_N(y_{i,1}|x(t_1,\tau))p_{t_0}^{t_1}(\tau)d\tau\times$$
$$\int_{t_0}^{t_1}\int_{t_1}^{t_2} f_N(y_{i,2}|x(t_2,\tau))f_N(y_{i,3}|x(t_3,\tau)+x^\star(t_3,\tau'))p_{t_0}^{t_1}(\tau)p_{t_1}^{t_2}(\tau')d\tau d\tau',$$

$$< 1,1,1 > : \mu_0\mu_{0\to 1}\mu_{1\to 2} \times \int_{t_0}^{t_1} f_N(y_{i,1}|x(t_1,\tau))p_{t_0}^{t_1}(\tau)d\tau\times$$
$$\int_{t_0}^{t_1}\int_{t_1}^{t_2} f_N(y_{i,2}|x(t_2,\tau)+x^\star(t_2,\tau'))p_{t_0}^{t_1}(\tau)p_{t_1}^{t_2}(\tau')d\tau d\tau'\times$$
$$\int_{t_0}^{t_1}\int_{t_1}^{t_2}\int_{t_2}^{t_3} f_N(y_{i,3}|x(t_3,\tau)+x^\star(t_3,\tau')+x^\star(t_3,\tau''))p_{t_0}^{t_1}(\tau)p_{t_1}^{t_2}(\tau')p_{t_2}^{t_3}(\tau'')d\tau d\tau' d\tau'',$$

where $\gamma$ is the baseline titer level in the kinetic model.

The key parameter of interest is the probability of infection $\mu$ between each round and for each participant. We can model this probability as:

$$\mu_i(t_a,t_b) = 1-\exp\left(-\lambda\int_{t_a}^{t_b}\beta(\tau)d\tau\right),$$

where $t_a$ and $t_b$ are the left and right time bounds of the period of interest for participant $i$ (for instance times of baseline and followup samples), $\lambda$ is a scaling rate parameter, and $\beta(t)$ is the time-varying force of cholera infection as described in the previous section.

###### S4.2.2 Additional source of infection

As we saw that there continued to be boosts in antibodies during periods where there were few cholera cases detected at clinics, we decided to consider an alternative model that allowed for an additional component of infection risk that is independent of the incidence of clinical disease. Specifically, we expand on the model above assuming there is a source of antibody boost-causing exposures/infections that is constant in time and independent of the incidence of the time-varying force of cholera infection described above. We denote the constant rate as $\lambda'$, and the probability of exposure/infection becomes:

$$\begin{aligned}\mu_i(t_a, t_b) &= 1 - \exp\left(-\lambda \int_{t_a}^{t_b} \beta(\tau)d\tau - \int_{t_a}^{t_b} \lambda' d\tau\right) \\ &= 1 - \exp\left(-\lambda \int_{t_a}^{t_b} \beta(\tau)d\tau - \lambda'(t_b - t_a)\right)\end{aligned}.$$

##### S4.2.3 Relative importance of constant vs. time-varying sources of infection

To quantify the relative importance of time-varying vs. constant force of infection we compute the ratio of marginal effects. For a given time period $[t_a, t_b]$, and let $x = \lambda \int_{t_a}^{t_b} \beta(\tau)d\tau$ and $y = \lambda'(t_b - t_a)$, the ratio of marginal effects is given by:

$$\begin{aligned}R(t_a, t_b) &= \frac{\frac{\partial \mu_i(t_a,t_b)}{\partial x}}{\frac{\partial \mu_i(t_a,t_b)}{\partial y}} \\ &= \frac{x\, exp(-(x+y))}{y\, exp(-(x+y))} \\ &= \frac{x}{y} = \frac{\lambda \int_{t_a}^{t_b} \beta(\tau)d\tau}{\lambda'(t_b - t_a)}.\end{aligned}$$

We compute the risk ratio over all time periods, and find that its value is of 0.01 (0.00-0.05), 0.97 (0.07-3.98), 0.75 (0.01-3.76) for below 5, 5-64 and above 65 respectively.

##### S4.2.4 Uncertainty in kinetic model parameters

The previous section assumes that all kinetic parameter models are known. In practice these are inferred with uncertainty in Jones et al. (2022). To account for uncertainty we marginalize out kinetic model parameters $\mathbf{\Theta}$ using samples from the posterior distribution in Jones et al. (2022). The probability of a given sequence of infections $Pr(< l, m, n > |\lambda)$ is therefore:

$$\begin{aligned}Pr(< l, m, n > |\lambda) =& \int_{\Omega} Pr(< l, m, n > |\lambda, \mathbf{\Theta})d\Theta \\ \approx& \frac{1}{J}\sum_{j=1}^{J} Pr(< l, m, n > |\lambda, \mathbf{\Theta}^j) \\ \mathbf{\Theta}^j \sim& f(\mathbf{\Theta}),\end{aligned}$$

where $f(\mathbf{\Theta})$ is the posterior distribution of parameter set $\mathbf{\Theta}$, and $\mathbf{\Theta}^j$ is a random draw from such a distribution. Random draws from this posterior distribution are shown below.

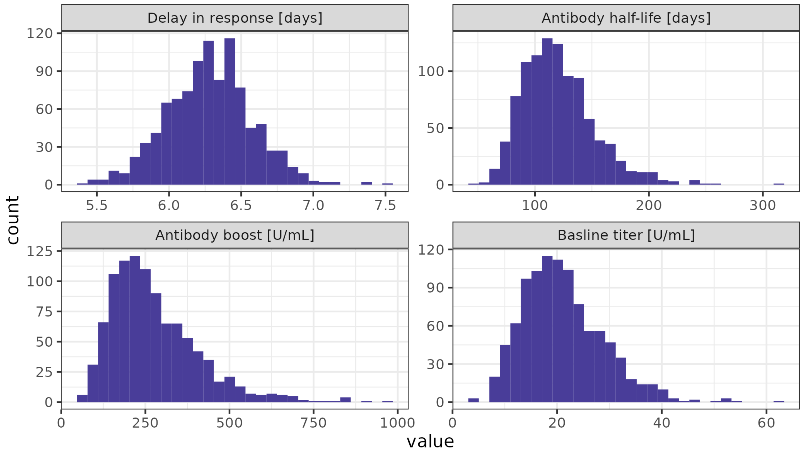


Figure S7: Posteriors of vibriocidal (Ogawa) antibody kinetics model parameters from Jones et al. 2022.

###### S4.2.5 Parameter pooling across age groups

Our final model infers the time-varying and constant seroincidence parameters jointly across the three age classes. Our final model implements pooling of both parameter values across age classes as:

$$\log \lambda_a \sim (\mu_{\log \lambda}, \sigma_{\log \lambda}),$$
$$\log \lambda'_a \sim (\mu_{\log \lambda'}, \sigma_{\log \lambda'}),$$

where $\mu$ and $\sigma$ are respectively the mean and the standard deviations of the time-varying and constant parameters on the log scale.

###### S4.2.6 Priors

We use the following priors in the final pooled sero-incidence model:

$$\mu_{\log \lambda} \sim \mathcal{N}(-4, 1),$$
$$\sigma_{\log \lambda} \sim \mathcal{N}(0, 1),$$
$$\mu_{\log \lambda'} \sim \mathcal{N}(-5, 1),$$
$$\sigma_{\log \lambda'} \sim \mathcal{N}(0, 1).$$

###### S4.2.7 Posterior retrodictive checks of serological trajectories

The following plot illustrates the titer trajectories for each participant classified by their most probable infection profile (panel/facet) along with the posterior probability of having that infection profile (color).

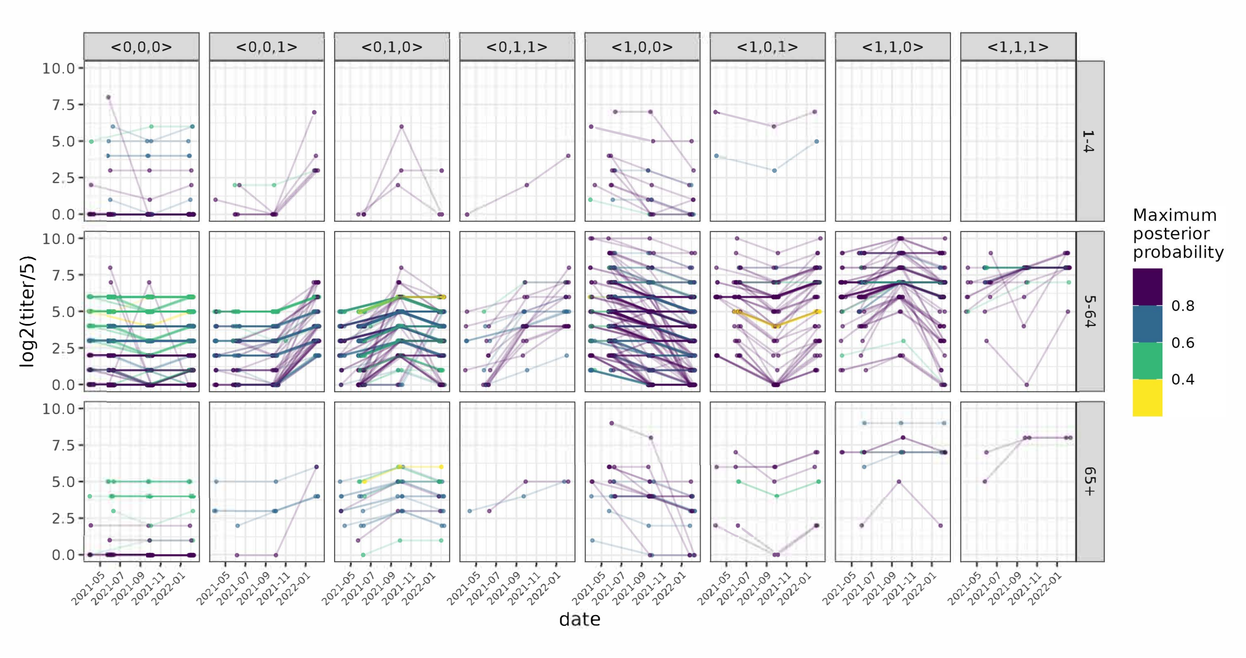


Figure S8: Posterior retrodictive checks of serological trajectories.

##### S4.2.8 Key posterior parameter draws

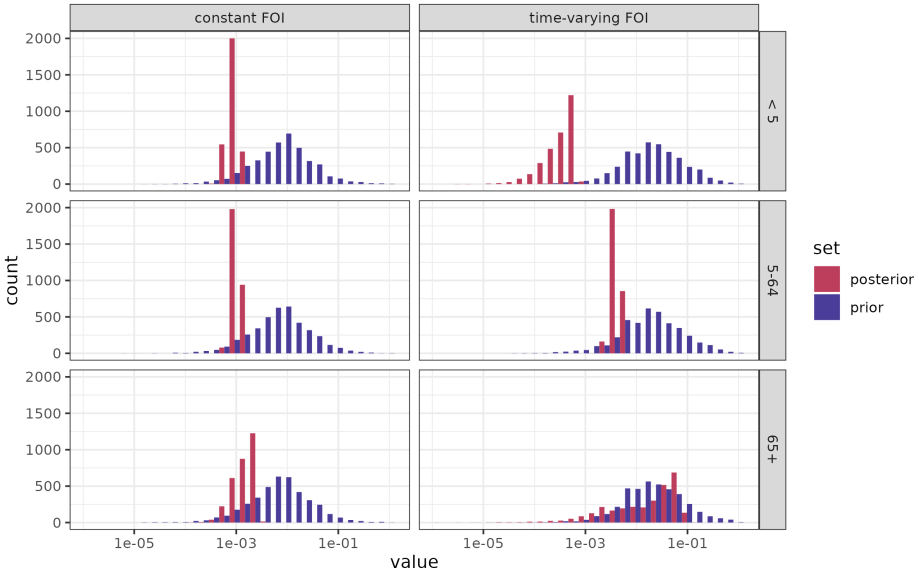


Figure S9: Sero-incidence model key parameter prior and posterior draws.

### S5 Medically-attended suspected case to true cholera case ratio

The ratio of suspected cases to the estimated true cholera incidence at health facilities decreases with age and is 10.5 (7.0-10.5) overall; for every 10.5 suspected cases that visit a health facility, 1 will be a true cholera case.

Table S2: Medically-attended suspected case to true cholera case ratio.

| Age group | Suspected case incidence per 1,000 | True cholera incidence per 1,000 | Ratio |
|---|---|---|---|
| 1-4 | 10.5 | 0.5 (0.4-0.7) | 21.0 (15.0-26.2) |
| 5-64 | 1.2 | 0.2 (0.1-0.2) | 6.0 (6.0-12.0) |
| 65+ | 1.9 | 0.5 (0.3-0.8) | 3.8 (2.4-6.3) |
| Overall | 2.1 | 0.2 (0.2-0.3) | 10.5 (7.0-10.5) |

Table S3: Number of individuals using different drinking water sources in week prior to survey for serological cohort (N=1,785; multiple sources allowed per individual)

| Serosurvey round | R1 | R2 | R3 |
|---|---|---|---|
| Season | Spring | Fall | Winter |
| Protected well (n) | 3 | 0 | 0 |
| Unprotected spring (n) | 3 | 0 | 0 |
| Rainwater (n) | 0 | 0 | 3 |
| Piped (n) | 254 | 112 | 86 |
| Tap (n) | 135 | 1 | 18 |
| Surface (n) | 0 | 0 | 101 |
| Tubewell (n) | 1403 | 1683 | 1685 |

#### S6 Drinking water sources

For the individuals in our serologic cohort (N=1,785), the use of piped and tap water as the primary water source in the week prior to the survey markedly reduced across seasons, from spring to winter, within the course of one year. Despite multiple water sources being permitted to be selected per individual, there was more piped and tap water in use during the high transmission season, which could be indicative of a lack of consistent water service and/or contamination.

#### S7 True symptomatic *V. cholerae* incidence for different definitions of diarrhea severity

Estimates of the true *V. cholerae* annualized symptomatic cholera incidence rates (using data from the community and clinics) per 1,000 population vary only slightly when using different definitions of diarrhea (mild, moderate, severe) to determine the probability of healthcare seeking. As the severity of diarrhea increases, our estimates of the true incidence of symptomatic cholera decreases.

Table S4: Estimates of annualized symptomatic incidence rates per 1,000 population

| Diarrhea type | 1-4 | 5-64 | 65+ | Overall |
|---|---|---|---|---|
| Mild | 2.7 (1.6-4.2) | 1.2 (1.0-1.5) | 4.6 (2.2-8.1) | 1.5 (1.2-1.8) |
| Moderate | 2.1 (1.3-3.0) | 0.7 (0.6-0.9) | 2.0 (1.0-3.4) | 0.9 (0.7-1.0) |
| Severe | 1.1 (0.7-1.6) | 0.4 (0.3-0.5) | 1.2 (0.6-1.9) | 0.5 (0.4-0.6) |
